# Tabular Foundation AI Models in Healthcare: A WHO-Inspired Benchmark for Responsible AI and Trustworthiness

**DOI:** 10.64898/2026.11.23.26368797

**Authors:** Kazi Sakib Hasan, Most. Afia Anjum Borsha, Raiseen Jahan Ritu, Lotifur Shabbir

## Abstract

Tabular foundation models (TFMs) are increasingly used for structured prediction, yet their suitability for healthcare applications remains uncertain because predictive performance alone does not establish fairness, probabilistic reliability, or robustness. We developed a multidimensional benchmark to compare four TFMs: TabFM, TabPFN, TabDPT, and TabICL, against Logistic Regression and XGBoost across a curated, standardized suite of ten WHO-aligned healthcare classification datasets across diverse disease domains. Additionally, we harmonized clinical schemas, standardized protected sex attributes and outcome targets, and released both balanced-scale and natural-prevalence variants of the datasets to establish a benchmarking resource in healthcare machine learning. All models were evaluated using identical five-fold stratified cross-validation partitions and standardized preprocessing. Performance was assessed using 20 metrics spanning predictive discrimination, sex-based fairness, probability calibration, and robustness to Gaussian feature perturbations. A composite Clinical Model Evaluation Score (CMES) was used to summarize performance across these dimensions. Google TabFM achieved the highest mean CMES (0.8811), followed closely by TabDPT (0.8796), TabICL (0.8775), TabPFN (0.8756), XGBoost (0.8650), and Logistic Regression (0.8549). The Friedman test indicated significant differences among models 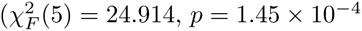; Kendall’s *W* = 0.498). However, post-hoc comparisons revealed no statistically significant differences among the four TFMs after Holm correction. TFMs generally demonstrated stronger discrimination and probabilistic calibration than conventional baselines, with TabFM achieving the highest mean Area Under the Receiver Operating Characteristic (0.9327) and the lowest expected calibration error (0.0674). Conversely, Logistic Regression exhibited the greatest stability under feature perturbations, while fairness performance varied substantially across datasets and models. These findings highlight the potential of TFMs for small-data healthcare prediction while demonstrating that their advantages are dimension-dependent. Multidimensional, dataset-specific evaluation and external validation remain essential before clinical implementation. All analysis codes, original datasets, and multiple variants of benchmark datasets are publicly available in a GitHub repository to ensure reproducibility and future research.

**Author Summary:** Tabular foundation AI models are increasingly used to make predictions from healthcare data, but their accuracy alone does not guarantee that they are fair, reliable, or robust. This study aimed to assess how well these models perform in disease prioritized by World Health Organization, and whether they can be trusted across different evaluation dimensions. The researchers compared four tabular foundation models (TabFM, TabPFN, TabDPT, and TabICL) with Logistic Regression and XGBoost using 10 healthcare datasets covering different disease areas. They evaluated predictive performance, fairness across sexes, reliability of predicted probabilities, and robustness to changes in input data. Google TabFM achieved the highest overall score, and foundation models generally performed better in prediction and probability calibration, while Logistic Regression was more stable under small input changes. The findings suggest that tabular foundation models could be promising for healthcare prediction, particularly when working with limited data. However, no model performed best across every dimension, and differences between foundation models were relatively small. Careful, dataset-specific evaluation and further validation in real-world clinical settings are needed before these models can be reliably used in healthcare.

## 1 Introduction

Tabular foundation models (TFMs) represent an emerging paradigm for structured data prediction, leveraging patterns learned across large pretraining corpora to generalize to new tasks with minimal or no dataset-specific training. Early progress was catalyzed by TabPFN, which demonstrated strong predictive performance on small tabular datasets through transformer-based tabular in-context learning (ICL) [1]. Building on this foundation, recent TFMs have extended tabular ICL through diverse architectural designs and pretraining strategies, including TabFM [2], TabICL [3], and TabDPT [4]. However, as these models gain attention for high-stakes applications such as digital health, strong accuracy on general-purpose tabular benchmarks does not by itself establish how they perform on clinical data or whether their probability estimates and failure modes are suitable for healthcare decision-making.

Many studies evaluated Machine Learning (ML) models on healthcare data, focusing on model’s discriminative power [5, 6]. However, evaluating machine learning models for healthcare requires assessing reliability beyond a single aggregate accuracy score. Predicted probabilities must be well calibrated so that estimated risk aligns with observed outcome frequencies, because poor calibration can distort clinical risk stratification and deteriorate across shifting patient populations [7, 8]. Fairness also requires explicit scrutiny because measurement disparities and historical clinical workflows can induce unequal error rates or selection rates across demographic subgroups such as biological sex [9, 10]. Furthermore, deployment settings routinely introduce recording variability and feature noise, meaning models should remain stable under input perturbations rather than exhibiting abrupt probability shifts or flipped classifications [11]. Contemporary clinical AI governance frameworks therefore emphasize multidimensional evaluation encompassing predictive discrimination, uncertainty calibration, algorithmic fairness, and robustness [12]. Conducting such an evaluation rigorously requires not only harmonized metrics, but also a disease-oriented dataset suite anchored in real global public health priorities.

The World Health Organization (WHO) Global Health Estimates provide an epidemiological basis for selecting conditions that reflect major global mortality and morbidity burdens. According to WHO estimates for 2023, five of the ten leading causes of death worldwide are ischaemic heart disease, stroke, chronic obstructive pulmonary disease (COPD), Alzheimer disease and other dementias, and diabetes mellitus [13]. Including clinical classification tasks for these five conditions anchors our benchmark in the leading drivers of global mortality, alongside cardiac arrhythmia, which is closely connected to ischaemic heart disease because myocardial ischaemia induces electrical instability and lethal ventricular arrhythmias that frequently precipitate sudden cardiac death [14]. To broaden coverage beyond leading causes of death to WHO global elimination and chronic morbidity priorities, the suite also incorporates hepatitis and chronic liver disease, reflecting WHO’s 2030 viral hepatitis elimination agenda in which chronic hepatitis B and C infections serve as major drivers of chronic liver disease, cirrhosis, and liver cancer. Finally, autism spectrum disorder (ASD) and thyroid disease represent high-burden neurodevelopmental and endocrine conditions characterized by pronounced, opposing sex asymmetries: ASD affects approximately 1 in 127 people globally with substantially higher prevalence and diagnostic rates in males than in females [15, 16], whereas thyroid disorders rank among the most prevalent endocrine conditions worldwide with markedly higher incidence and prevalence in women than in men [17]. Together, these ten tasks span distinct organ systems, clinical modalities, and demographic risk profiles.

Evaluating TFMs across this multi-disease suite makes it possible to examine whether general-purpose tabular ICL models capture clinically meaningful patterns consistently across heterogeneous healthcare domains. Because these datasets encompass demographic variables, clinical symptoms, physiological measurements, and biochemical markers, comparing models across all ten tasks reveals whether strong performance remains consistent beyond isolated disease areas. At the same time, high performance on one evaluation dimension does not imply adequacy on another: a model with high discrimination may still produce miscalibrated probabilities, exhibit disparity between male and female patients, or react sensitively to minor input noise. Moreover, apparent associations or strong cross-validation scores within curated datasets should not be interpreted as evidence of causal mechanisms or autonomous diagnostic readiness. Rather, a multidimensional benchmark clarifies specific operational strengths and trade-offs across candidate TFMs before any downstream clinical adaptation is considered.

Previous evaluations of TFMs have focused primarily on broad multi-domain leaderboards and headline predictive accuracy or AUROC, leaving open how leading tabular ICL architectures compare under unified clinical trustworthiness criteria. Comparing TabFM [2], TabPFN [1], TabDPT [4], and TabICL [3] independently across disparate preprocessing pipelines risks conflating model behavior with differences in data splitting, imputation, or metric definitions. A fair head-to-head comparison therefore requires evaluating all four models on identical stratified cross-validation partitions, applying missing-value imputation strictly within each training fold to prevent data leakage, and enforcing uniform positive-class definitions, protected-group mappings, and decision thresholds. Standardizing these experimental controls ensures that observed differences across fairness, discrimination, calibration, and robustness reflect intrinsic model properties rather than evaluation artifacts.

A rigorous multidimensional benchmark should evaluate complementary metric families that answer distinct clinical and statistical questions. Predictive discrimination metrics, including the area under the receiver operating characteristic curve (AUROC), sensitivity, specificity, positive predictive value (PPV), and negative predictive value (NPV) measure how effectively a model separates positive and negative outcomes. Uncertainty calibration metrics like Expected Calibration Error (ECE), negative log-likelihood (NLL), Brier score, calibration slope, and calibration intercept assess whether predicted positive probabilities correspond to observed event frequencies and remain free from systematic over- or under-confidence [7]. Group fairness metrics, such as Disparate Impact (DI), Equal Opportunity Difference (EOD), Demographic Parity Difference (DPD), Equalized Odds Difference (EqOD), and Predictive Parity Difference (PPD) quantify prediction equity between male and female subgroups [10]. Finally, robustness metrics under controlled Gaussian feature perturbations, signed degradations in AUROC, accuracy, and F1-score, alongside prediction flip rate and mean absolute probability change (MAPC) capture how reliably individual predictions and overall performance hold under measurement noise [12]. Assessing all 20 metrics jointly provides a comprehensive profile of model trustworthiness rather than relying on a single leaderboard ranking.

Against this background, benchmarking TabFM, TabPFN, TabDPT, and TabICL across ten WHO-aligned healthcare classification datasets establishes empirical evidence on the relative trade-offs of modern tabular ICL models in clinical settings. The objective is not to replace domain-specific clinical validation or establish readiness for autonomous medical deployment, which would require external validation in prospective target populations [7, 8]. Instead, a standardized internal cross-validation benchmark provides a reproducible baseline identifying which TFMs balance predictive discrimination, sex-based fairness, probabilistic calibration, and noise resilience across diverse healthcare tasks, and where specific vulnerabilities warrant targeted mitigation.

Accordingly, this study investigates how TabFM, TabPFN, TabDPT, and TabICL compare across diverse tabular healthcare classification datasets by addressing six research questions:

1. **Fairness and equity:** How do the four TFMs compare across sex-based fairness metrics (DI, EOD, DPD, EqOD, and PPD) when evaluated on identical clinical partitions and fixed classification thresholds?
2. **Predictive discrimination:** How effectively do TFMs distinguish positive and negative clinical outcomes across ten WHO-aligned healthcare datasets in terms of AUROC, sensitivity, specificity, PPV, and NPV?
3. **Uncertainty calibration:** How reliable are the models’ positive-class probability estimates, as measured by ECE, NLL, Brier score, calibration slope, and calibration intercept?
4. **Perturbation robustness:** To what extent do model predictions, probabilities, and discrimination metrics (e.g., AUROC, F1-score) remain stable when test features (excluding protected sex and outcome labels) are subjected to standardized Gaussian noise?
5. **Baseline comparison:** How do the four TFMs perform against traditional ML baselines (Logistic Regression, XGBoost) in terms of fairness, predictive power, uncertainty calibration, and perturbation robustness?
6. **Composite evaluation:** How do the baseline models and TFMs compare in terms of an overall composite score integrating dimensions of predictive discrimination, positive-class detection, fairness, probabilistic calibration, and robustness, as quantified by DI, DPD, AUROC, sensitivity, Brier score, ECE, AUROC degradation, and F1 degradation?

### 1.1 Related Work

For tabular prediction tasks, gradient-boosted decision trees (GBDTs) long served as the default baseline over deep neural architectures. In a large-scale empirical comparison across 176 classification datasets, McElfresh et al. demonstrated that neither GBDTs nor task-specific neural networks dominated universally, showing instead that dataset metafeatures and tuning budgets heavily governed model rankings [18]. To eliminate per-dataset gradient optimization altogether, Hollmann et al. introduced TabPFN, a transformer pretrained on millions of synthetic causal datasets to approximate Bayesian posterior predictive distributions via tabular in-context ICL in a single forward pass [19]. While this initial formulation was constrained to small, purely numerical tables without missing entries, a subsequent iteration (TabPFNv2) expanded the synthetic structural causal prior to accommodate missing values, categorical variables, and heterogeneous feature distributions [1], with later scaling efforts extending context capacity and features [20].

Concurrently, competing TFMs have explored distinct pretraining paradigms and scalable attention mechanisms. Qu et al. proposed TabICL, employing a two-stage column-then-row attention architecture that embeds rows independently before performing cross-row ICL on large datasets [3], and later introduced TabICLv2 with scalable softmax attention and diversified synthetic data generation [21]. Departing from purely synthetic priors, Ma et al. developed TabDPT, which combines retrieval-augmented in-context inference with self-supervised pretraining on real-world tabular corpora [4]. Most recently, Google Research introduced TabFM to strengthen zero-shot generalization across heterogeneous tabular schemas [2]. To standardize comparisons across these rapidly evolving architectures, Erickson et al. established the TabArena living benchmark over 51 curated datasets, showing that modern tabular ICL models now match or surpass heavily tuned GBDT ensembles on independent and identically distributed (IID) tasks [22]. However, a critical synthesis by Kurashkin et al. cautioned that benchmark gains on general independent and identically distributed (IID) leaderboards do not resolve open questions regarding conditional probability coverage, distribution-shift resilience, and fairness [23], underscoring the need to evaluate TFMs within domain-specific, high-stakes settings such as digital healthcare.

#### 1.1.1 Tabular Foundation Models in Clinical and Healthcare Domain

Transitioning tabular prediction models into healthcare introduces stricter methodological and clinical requirements than general-purpose benchmarking. The TRIPOD+AI consensus statement updated clinical prediction reporting standards to cover ML alongside regression, emphasizing probability calibration, and subgroup evaluation [24]. Similarly, the FUTURE-AI guideline formalized six core principles: fairness, universality, traceability, usability, robustness, and explainability, as prerequisites for trustworthy medical AI [12]. Against these standards, recent empirical studies have begun testing TFMs on specialized medical tasks. Zhang et al. evaluated TabPFN against five conventional ML algorithms in a retrospective cohort of 986 neonates for hyperbilirubinemia risk stratification, reporting strong discrimination, Brier scores, and net clinical benefit across both internal cross-validation and temporal holdout cohorts [25]. In musculoskeletal screening, Liang and Wang combined anatomical pose features with TabPFN and Shapley-based explanations to classify forward head posture across 1,577 adolescents [26]. Beyond physiological screening, Shawosh et al. evaluated TabPFN for explainable health insurance claim prediction, highlighting its utility in reducing dataset-specific hyperparameter tuning overhead [27].

Despite encouraging early results, the existing healthcare literature on TFMs exhibits two notable limitations. First, applied clinical evaluations to date have focused almost exclusively on TabPFN as a standalone representative of tabular ICL, comparing it against classical baselines such as XGBoost or logistic regression rather than against competing TFMs like TabFM, TabDPT, or TabICL. Second, prior medical evaluations typically examine a single clinical endpoint or institutional cohort, making it difficult to assess whether observed strengths generalize across diverse disease categories, sample sizes, and feature modalities. Consequently, there remains little comparative evidence on how distinct TFMs perform across a standardized, multi-disease public health benchmark.

#### 1.1.2 Trustworthiness Dimensions: Calibration, Fairness, and Perturbation Robustness

Beyond predictive discrimination, clinical usefulness depends on probabilistic calibration, demographic fairness, and stability under noisy inputs [28]. Van Calster et al. emphasized that discrimination alone cannot guarantee safe risk estimation, as miscalibrated probabilities directly distort clinical decision thresholds [7], while Youssef et al. argued that calibration drift under shifting clinical environments necessitates recurring local validation [8]. Several recent studies have begun examining TFM calibration explicitly. Helli et al. demonstrated that standard TabPFN can experience degraded accuracy and ECE under temporal distribution shifts, motivating a drift-resilient causal prior to preserve calibrated predictions [29]. Extending this inquiry to multimodal embeddings, Zhang et al. reported that TabPFN consistently lowered NLL and ECE relative to classical classification heads across 22,820 evaluation episodes, though its relative accuracy advantage narrowed under high feature dimensionality and scarce labeled contexts [30]. Whether these calibration properties hold uniformly across TabFM, TabDPT, and TabICL on clinical tabular data, when assessed with logistic calibration slope and intercept alongside ECE, NLL, and Brier score remains largely unexamined.

Algorithmic fairness represents a parallel concern in healthcare AI, where biased sampling and unequal access to care can yield disparate error rates across protected attributes such as biological sex [10, 31, 32]. Addressing causal bias in prior-data fitted networks, Robertson et al. introduced FairPFN, demonstrating that standard synthetic-prior pretraining can propagate observational correlations tied to protected attributes unless counterfactual fairness is explicitly encoded during pretraining [33]. Most directly relevant to multi-model TFM comparison, Kenfack et al. conducted the first systematic investigation of group fairness across TabPFNv2, TabICL, and TabDPT, revealing that all three models exhibit measurable demographic parity, equalized odds, and equal opportunity gaps under standard context selection unless uncertainty-based or correlation-removal interventions are applied [34]. However, their benchmark evaluated general tabular datasets rather than clinical disease tasks, and did not include Google TabFM or pair fairness audits with calibration and noise-robustness profiling.

Finally, real-world clinical measurements are routinely affected by sensor noise, transcription errors, and physiological variability, making perturbation robustness essential for reliable deployment. Djilani et al. investigated test-time adversarial robustness across TabPFN and TabICL on finance, cybersecurity, and healthcare datasets, showing that small, structured perturbations to test inputs with a fixed training context can significantly degrade accuracy and flip predictions [35]. Yet while targeted adversarial attacks establish worst-case vulnerability, less is known about how competing TFMs, including TabFM and TabDPT respond to standardized, non-adversarial Gaussian measurement noise across clinical covariates when protected demographic attributes remain unperturbed.

#### 1.1.3 Synthesis and Research Gap

Synthesizing recent advances from 2023 to 2026 reveals a clear disconnect between rapid architectural progress in TFMs and the multidimensional evidence required for clinical AI adoption. General-domain benchmarks such as TabArena establish strong predictive accuracy across TFMs [22], yet critical reviews note persistent gaps in uncertainty coverage, fairness, and shift robustness [23]. Meanwhile, emerging clinical studies test TabPFN in isolated single-disease cohorts [25, 26], and specialized trustworthiness studies examine fairness [34], distribution-shift calibration [29], or test-time perturbations [35] in isolation across non-standardized datasets. No prior study has conducted a controlled, head-to-head comparison of TabFM, TabPFN, TabDPT, and TabICL across a WHO-aligned suite of healthcare classification datasets a harmonized 20-metric framework spanning fairness, predictive discrimination, calibration, and Gaussian noise robustness. The present benchmark directly addresses this gap.

## 2 Materials and Methods

We establish a standardized, reproducible benchmarking pipeline across ten diverse healthcare classification tasks to systematically evaluate the reliability of emerging TFMs in clinical settings. Our experimental protocol compares four TFMs and two traditional ML models under identical data partitions, preprocessing rules, and evaluation criteria spanning four complementary dimensions: predictive discrimination, group fairness, uncertainty calibration, and robustness to input perturbations.

### 2.1 Benchmark Datasets

The benchmark suite comprises ten publicly available tabular healthcare classification datasets selected to represent WHO-prioritized global mortality and chronic morbidity conditions across diverse clinical domains, organ systems, and demographic risk profiles. The study is designed to comparatively evaluate TFMs and conventional ML classifiers under a constrained sample-size regime, with particular emphasis on predictive performance, fairness, calibration, and robustness across heterogeneous healthcare classification tasks. Table 1 summarizes the clinical domains, class distributions, and sample sizes of the selected datasets. To establish a small-scale evaluation setting and limit dataset-size disparities, datasets exceeding 700 observations were subjected to random majority-class downsampling, restricting the maximum sample size to 700 observations. This constraint is motivated by the small-data setting in which TFMs are particularly relevant, allowing their comparative performance to be examined against traditional ML approaches without relying on large task-specific datasets [3, 36]. This choice of 700 samples is consistent with previous empirical studies suggesting that ML models often require several hundred to over a thousand observations to achieve stable predictive performance [37, 38]. Accordingly, this benchmark investigates comparative model behavior under constrained data availability rather than establishing clinical generalizability, population-level risk estimation, or deployment readiness. The original dataset sources, processed datasets, and analysis code are made publicly available through the study’s GitHub repository to facilitate reproducibility [39]. Additionally, three supplementary datasets are made available through the GitHub repository, including a dataset with original sample size and natural class distribution, providing researchers with access to unaltered data for further investigation, alternative benchmarking configurations, and future comparative studies beyond the constraints of the present experimental setting [39]. We accessed the datasets for research purpose in October 2026, and had no access to information that could identify individual participants during or after data collection.

**Table 1.**
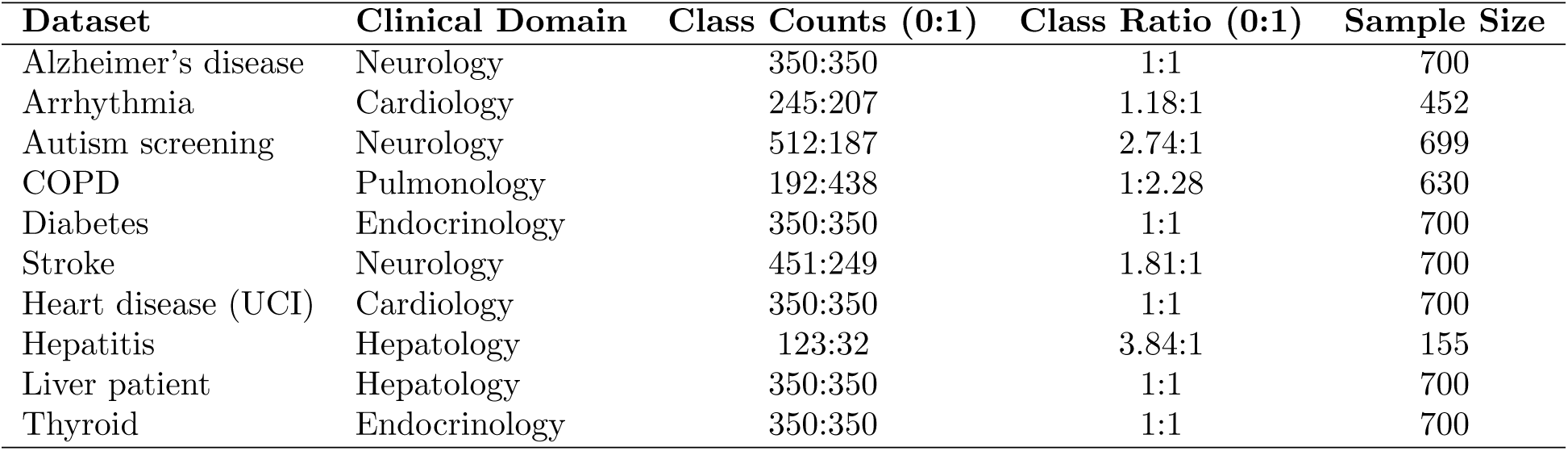
Clinical domains and class distributions of the datasets used in the study.

### 2.2 Data Preprocessing and Partitioning

All ten datasets undergo a uniform preparation pipeline prior to model evaluation in order to ensure cross-model comparability and eliminate preprocessing discrepancies.

1. **Standardized Protected Attribute and Target Encoding:** Across all datasets, the protected attribute (`sex`) is standardized as a binary variable with Female = 0 and Male = 1, and positioned as the first feature column. All classification tasks are formulated as binary prediction problems where the target variable (`outcome`), positioned as the final column, maps the negative class to 0 and the positive clinical class to 1. For datasets originally containing multi-class outcomes (e.g., Thyroid and Heart Disease UCI), all non-positive categories are consolidated into the negative class (0), while ensuring that intermediate risk categories are not improperly grouped into the negative class.
2. **Categorical Feature Encoding:** Non-numerical categorical attributes are mapped to integer representations via explicit manual label replacement (Python .replace()) rather than one-hot encoding. This design preserves the original tabular schema, prevents dimensionality inflation, and retains feature-level interpretability.
3. **Class Balancing:** Class distributions across all datasets are approximately balanced in some datasets due to random downsampling. For some datasets with sample size already lower than 700, their natural class ratio is preserved. All the balanced and original imbalanced dataset versions are archived in the study’s GitHub repository for reproducibility and future research.
4. **Cross-Validation and Leak-Free Imputation:** Each dataset is evaluated using 5-fold stratified cross-validation, preserving the approximate class distribution within every fold. Identical train/test fold assignments are enforced across all evaluated models. Missing values are retained in the prepared datasets and imputed independently within each cross-validation fold using *k*-nearest neighbors (KNN) imputation fit solely on the training fold, and subsequently applied to the corresponding test fold, thereby preventing data leakage.

### 2.3 Tabular Foundation Models

We benchmark four representative TFMs using their fixed, reproducible release versions: **TabFM** (`v1.0.0`) by Google, **TabPFN** (`v2.0`) by Prior Labs, **TabDPT** (`v1.3`) by Layer 6 AI, and **TabICL** (`v2`) by Inria. For each fold, every model is provided the identical training partition as context (training data) and outputs both continuous positive-class probabilities and discrete binary predictions on the held-out test partition. Binary predictions are generated using a fixed classification threshold of *τ* = 0.50 across all models as well as both clean and noisy test evaluations. Threshold tuning on the test set is strictly prohibited.

### 2.4 Baseline Models

We include two classical ML baselines: **Logistic Regression** (abbreviated as LogReg in several figures and tables) and **XGBoost**, to contextualize the performance of TFMs against established clinical modeling approaches. Both baselines are evaluated across the identical 5-fold stratified cross-validation as the foundation AI models. For Logistic Regression, input features are standardized prior to model fitting with the Stochastic Average Gradient Augmented (SAGA) optimizer. XGBoost is trained directly on the imputed features using the logistic loss objective.

Hyperparameter tuning is performed independently within each outer training fold via nested Bayesian optimization with a Tree-structured Parzen Estimator (TPE). For each model and outer fold, the optimizer maximizes the mean AUROC across shuffled inner stratified cross-validation splits of the outer training fold. KNN imputation (and feature standardization for Logistic Regression) is fit independently on each inner training split to strictly prevent data leakage during training. The complete hyperparameter search spaces are detailed in Table 2.

**Table 2.**
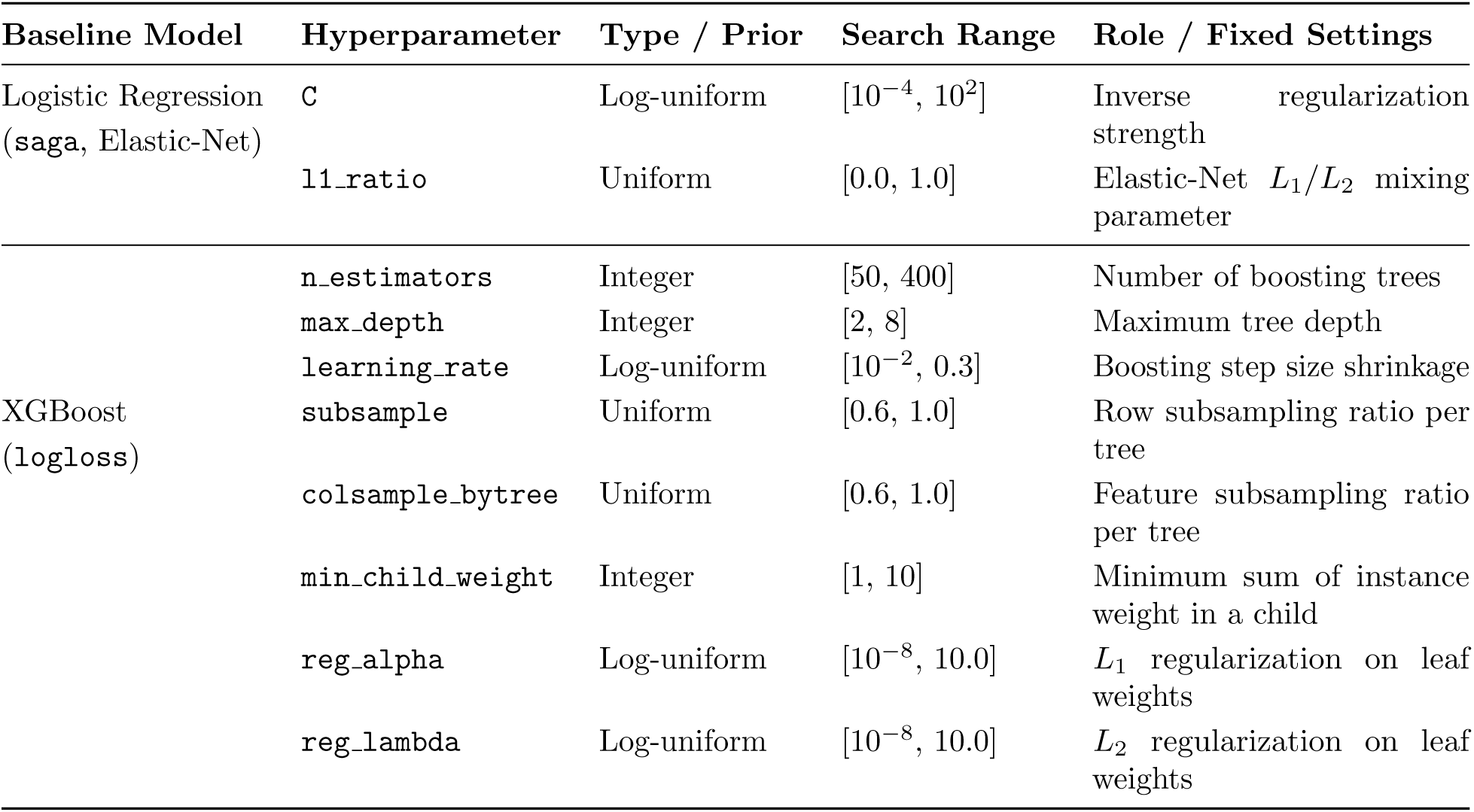
Hyperparameter search spaces for baseline models tuned via Bayesian optimization with a TPE.

| Baseline Model | Hyperparameter | Type / Prior | Search Range | Role / Fixed Settings |
|---|---|---|---|---|
| Logistic Regression (saga, Elastic-Net) | C | Log-uniform | [10<sup>−4</sup>, 10<sup>2</sup>] | Inverse regularization strength |
|  | l1_ratio | Uniform | [0.0, 1.0] | Elastic-Net *L*<sub>1</sub>/*L*<sub>2</sub> mixing parameter |
| XGBoost (logloss) | n_estimators | Integer | [50, 400] | Number of boosting trees |
|  | max_depth | Integer | [2, 8] | Maximum tree depth |
|  | learning_rate | Log-uniform | [10<sup>−2</sup>, 0.3] | Boosting step size shrinkage |
|  | subsample | Uniform | [0.6, 1.0] | Row subsampling ratio per tree |
|  | colsample_bytree | Uniform | [0.6, 1.0] | Feature subsampling ratio per tree |
|  | min_child_weight | Integer | [1, 10] | Minimum sum of instance weight in a child |
|  | reg_alpha | Log-uniform | [10<sup>−8</sup>, 10.0] | *L*<sub>1</sub> regularization on leaf weights |
|  | reg_lambda | Log-uniform | [10<sup>−8</sup>, 10.0] | *L*<sub>2</sub> regularization on leaf weights |

### 2.5 Evaluation Dimensions and Metrics

Model performance is assessed across 20 standardized metrics evenly distributed across four evaluation dimensions (5 metrics per dimension). All 20 metrics are computed independently on each held-out fold (*k* = 1*,…,* 5). For each dataset and model, fold-level results are aggregated across the 5 folds using the arithmetic mean and the sample standard deviation. Table 3 provides an overview of all benchmark metrics, their mathematical definitions, value ranges, and optimization directions.

**Table 3.**
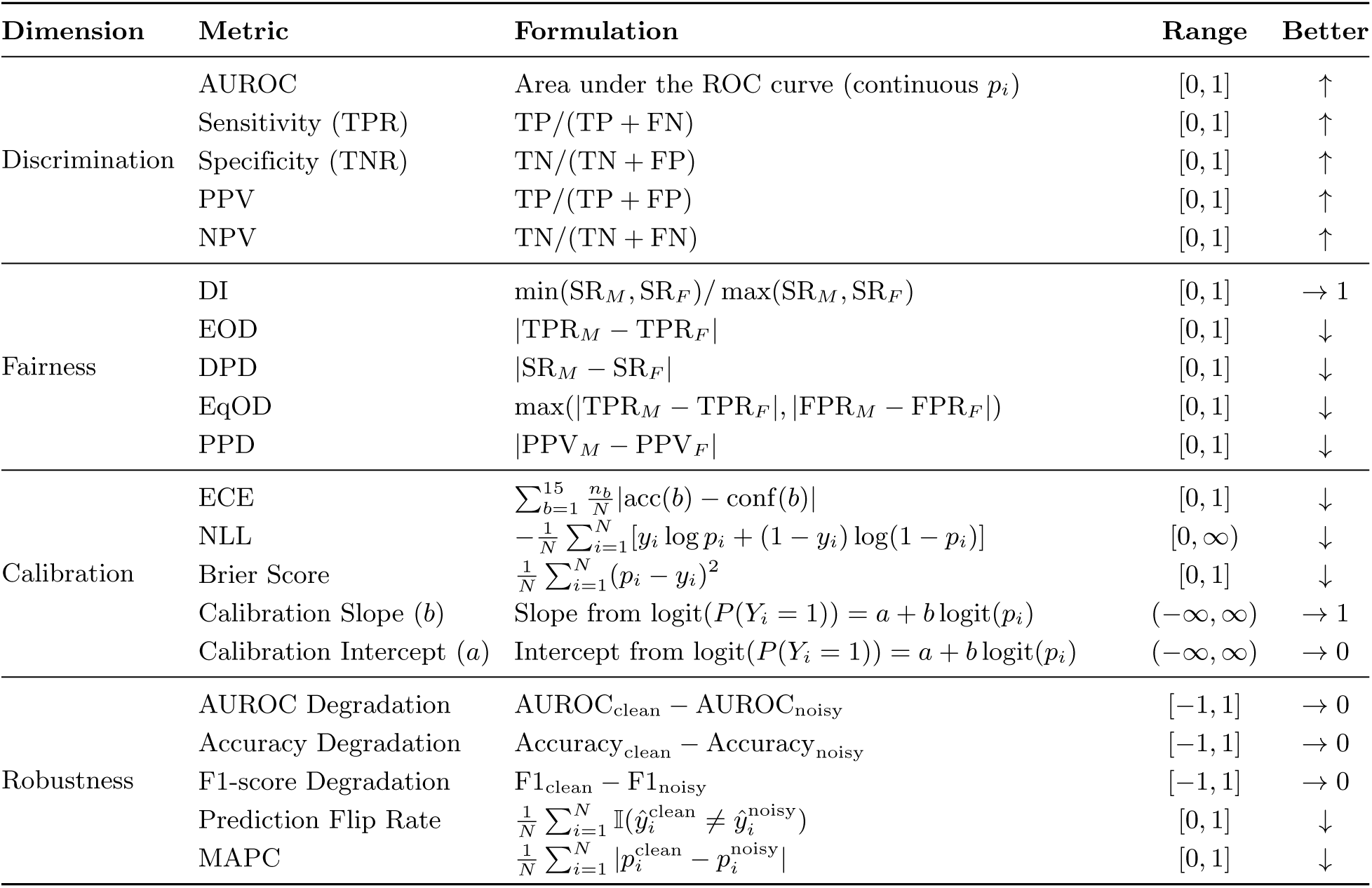
Summary of the 20 benchmark evaluation metrics across four dimensions.

| Dimension | Metric | Formulation | Range | Better |
|---|---|---|---|---|
| Discrimination | AUROC | Area under the ROC curve (continuous *p*<sub>*i*</sub>) | [0, 1] | ↑ |
|  | Sensitivity (TPR) | TP/(TP + FN) | [0, 1] | ↑ |
|  | Specificity (TNR) | TN/(TN + FP) | [0, 1] | ↑ |
|  | PPV | TP/(TP + FP) | [0, 1] | ↑ |
|  | NPV | TN/(TN + FN) | [0, 1] | ↑ |
| Fairness | DI | min(SR<sub>*M*</sub>, SR<sub>*F*</sub>)/ max(SR<sub>*M*</sub>, SR<sub>*F*</sub>) | [0, 1] | → 1 |
|  | EOD | \|TPR<sub>*M*</sub> − TPR<sub>*F*</sub>\| | [0, 1] | ↓ |
|  | DPD | \|SR<sub>*M*</sub> − SR<sub>*F*</sub>\| | [0, 1] | ↓ |
|  | EqOD | max(\|TPR<sub>*M*</sub> − TPR<sub>*F*</sub>\|, \|FPR<sub>*M*</sub> − FPR<sub>*F*</sub>\|) | [0, 1] | ↓ |
|  | PPD | \|PPV<sub>*M*</sub> − PPV<sub>*F*</sub>\| | [0, 1] | ↓ |
| Calibration | ECE | $\sum_{b=1}^{15} \frac{n_b}{N} \lvert \mathrm{acc}(b) - \mathrm{conf}(b) \rvert$ | [0, 1] | ↓ |
|  | NLL | $-\frac{1}{N}\sum_{i=1}^{N}[y_i \log p_i + (1-y_i)\log(1-p_i)]$ | [0, ∞) | ↓ |
|  | Brier Score | $\frac{1}{N}\sum_{i=1}^{N}(p_i - y_i)^2$ | [0, 1] | ↓ |
|  | Calibration Slope (*b*) | Slope from logit(*P*(*Y*<sub>*i*</sub> = 1)) = *a* + *b* logit(*p*<sub>*i*</sub>) | (−∞, ∞) | → 1 |
|  | Calibration Intercept (*a*) | Intercept from logit(*P*(*Y*<sub>*i*</sub> = 1)) = *a* + *b* logit(*p*<sub>*i*</sub>) | (−∞, ∞) | → 0 |
| Robustness | AUROC Degradation | AUROC<sub>clean</sub> − AUROC<sub>noisy</sub> | [−1, 1] | → 0 |
|  | Accuracy Degradation | Accuracy<sub>clean</sub> − Accuracy<sub>noisy</sub> | [−1, 1] | → 0 |
|  | F1-score Degradation | F1<sub>clean</sub> − F1<sub>noisy</sub> | [−1, 1] | → 0 |
|  | Prediction Flip Rate | $\frac{1}{N}\sum_{i=1}^{N} \mathbb{I}(\hat{y}_i^{\mathrm{clean}} \neq \hat{y}_i^{\mathrm{noisy}})$ | [0, 1] | ↓ |
|  | MAPC | $\frac{1}{N}\sum_{i=1}^{N} \lvert p_i^{\mathrm{clean}} - p_i^{\mathrm{noisy}} \rvert$ | [0, 1] | ↓ |

#### 2.5.1 Predictive Discrimination

Discrimination metrics assess each model’s capacity to distinguish between positive (*y_i_* = 1) and negative (*y_i_* = 0) clinical outcomes. Using the confusion matrix counts (TP, TN, FP, FN) derived from thresholded predictions *ŷ_i_*, we compute Sensitivity, Specificity, PPV, and NPV. The AUROC is computed from the continuous positive-class probabilities *p_i_*.

#### 2.5.2 Fairness and Equity

Group fairness is evaluated with respect to biological sex, comparing predictions between Male (*M*, encoded as 1) and Female (*F*, encoded as 0) subgroups under the identical positive-class definition (*y* = 1). For each subgroup *g ∈ {M, F}*, we compute the selection rate SR*_g_* = *P* (*ŷ* = 1 *| g*), true positive rate TPR*_g_* = TP*_g_/*(TP*_g_* + FN*_g_*), false positive rate FPR*_g_* = FP*_g_/*(FP*_g_* + TN*_g_*), and positive predictive value PPV*_g_* = TP*_g_/*(TP*_g_* + FP*_g_*). From these group-level rates, we calculate DI, EOD, DPD, EqOD, and PPD as defined in Table 3.

#### 2.5.3 Uncertainty and Calibration

Calibration evaluates whether predicted positive-class probabilities *p_i_* accurately reflect empirical event frequencies. Prior to logarithmic or logit transformations, all predicted probabilities are clipped to [10*^−^*^15^, 1 *−* 10*^−^*^15^] to ensure numerical stability. ECE is calculated by partitioning predictions into 15 equal-width probability bins covering [0, 1] (with the final bin inclusive of *p_i_* = 1), computing the sample-weighted absolute difference between mean observed outcome acc(*b*) and mean predicted confidence conf(*b*) across bins containing *n_b_* observations out of *N* total test samples. In addition, we compute NLL and the Brier Score. Finally, Calibration Intercept (*a*, ideal value 0) and Calibration Slope (*b*, ideal value 1) are estimated on the test-fold observations via logistic calibration regression:

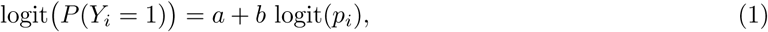

#### 2.5.4 Robustness under Feature Perturbation

To quantify sensitivity to noise and input perturbations, we inject zero-mean additive Gaussian noise into the test set of each cross-validation fold while keeping the context data, the protected attribute (`sex`), and the ground-truth target labels (`outcome`) completely unchanged. For each observation *i* and each remaining input feature *j*, the perturbed test value *x^′^_ij_* is generated as:

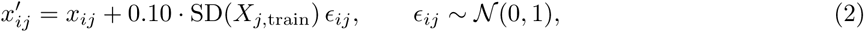

where SD(*X_j,_*_train_) denotes the sample standard deviation of feature *j* computed exclusively from the current fold’s training set. The perturbation magnitude was set to 10% (0.10) of the training-fold feature standard deviation to introduce a controlled, relatively mild disturbance while preserving the original feature scale. This choice is consistent with prior robustness-testing studies that investigate classifier sensitivity under feature-scaled Gaussian noise and systematically evaluate noise severity using incremental perturbation levels [40, 41].

Each fitted model is evaluated on both the clean test fold and its noisy counterpart to compute five robustness metrics: performance degradations in AUROC, Accuracy, and F1-score (*D_M_* = *M*_clean_ *− M*_noisy_), Prediction Flip Rate, and MAPC. For degradation metrics *D_M_*, positive values indicate performance deterioration under noise, zero indicates invariance, and negative values indicate slight performance improvement; negative values are preserved verbatim without truncation to zero or absolute-value transformation, such that values closer to zero consistently signify superior stability.

### 2.6 Composite Metrics and Statistical Analysis

A composite evaluation is constructed using eight metrics selected to capture complementary dimensions of model performance relevant to the objectives of this study: DI, DPD, AUROC, Sensitivity, Brier Score, ECE, AUROC Degradation, and F1 Degradation. This selection prioritizes predictive discrimination, positive-class detection, fairness in group-level outcomes, probabilistic accuracy and calibration, and robustness to input perturbations. Specifically, AUROC and Sensitivity characterize discriminative ability and positive-class detection, respectively, while DI and DPD capture disparities in selection rates across biological sex subgroups. The Brier Score and ECE jointly assess the quality of probabilistic predictions, reflecting both prediction error and calibration reliability. AUROC Degradation and F1 Degradation further quantify the extent to which discriminative performance deteriorates under feature perturbations, providing complementary perspectives on model robustness. These metrics are therefore selected to align the composite evaluation with the central research objectives, rather than to exhaustively represent all 20 metrics. The remaining metrics are retained for individual analyses to provide more granular assessments of specific performance characteristics but are not incorporated into the composite score to avoid broadening its scope beyond the dimensions of primary interest. The composite scores were calculated using the mean values of the selected evaluation metrics across the five-fold cross-validation procedure.

#### 2.6.1 Composite Metric Construction

Four dimension-specific composite scores were constructed to provide a unified representation of model performance. Each metric was transformed, where necessary, so that higher values consistently indicated better performance. The component metrics within each dimension were assigned equal weights.

##### Fairness Composite Score (FCS)

Fairness was assessed using DI and DPD. DI measures the ratio of positive prediction rates between protected and reference groups, with values closer to 1 indicating greater parity. DPD measures the absolute difference in positive prediction rates between groups, for which lower values indicate better demographic parity. Accordingly, the FCS was defined as:

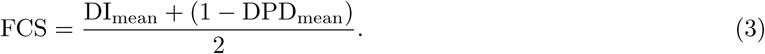

The transformation (1 *−* DPD_mean_) converts DPD into a higher-is-better measure, allowing both fairness metrics to contribute consistently to the composite score.

##### Predictive Performance Composite Score (PPCS)

Predictive performance was evaluated using AUROC and Sensitivity. AUROC measures the model’s ability to discriminate between positive and negative classes across classification thresholds, whereas Sensitivity measures its ability to correctly identify positive cases. Both metrics range from 0 to 1, with higher values indicating better predictive performance. The corresponding composite score was calculated as:

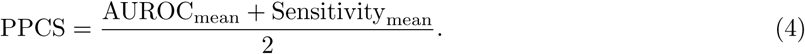

These metrics were selected to represent complementary aspects of clinical prediction: overall discrimination and the identification of positive cases.

##### Probabilistic Calibration Composite Score (PCCS)

Calibration was assessed using the Brier Score and ECE. The Brier Score quantifies the mean squared difference between predicted probabilities and observed outcomes, while ECE measures the discrepancy between predicted confidence and empirical accuracy across probability bins. Both metrics range from 0 to 1, with lower values indicating better calibration. To ensure a consistent higher-is-better direction, both metrics were inverted before aggregation:

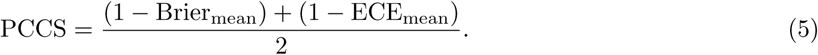

The inclusion of both metrics provides complementary information about probabilistic reliability, capturing overall probability error and the agreement between predicted confidence and observed frequencies.

##### Robustness Composite Score (RCS)

Robustness was evaluated using AUROC degradation (AUROC_Deg) and F1-score degradation (F1_Deg), which quantify changes in predictive performance under the specified degradation conditions. Since values closer to zero indicate smaller performance changes and therefore greater robustness, the absolute values of both degradation metrics were used. The RCS was defined as:

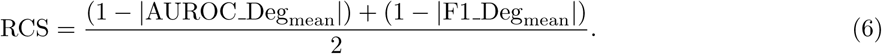

The absolute-value transformation ensures that both positive and negative deviations from zero are treated as changes in performance, rather than interpreting one direction of deviation as inherently more desirable. This formulation therefore rewards models with smaller absolute performance changes under degradation.

##### Clinical Model Evaluation Score (CMES)

The four dimension-specific composite scores were combined into a final Clinical Model Evaluation Score (CMES) to obtain a single overall measure of model performance:

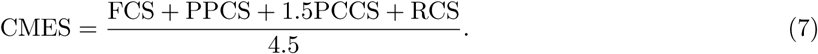

The four dimensions were not assigned identical weights. Fairness, predictive performance, and robustness each received a weight of 1, whereas probabilistic calibration received a weight of 1.5. This weighting scheme was specified to assign additional emphasis to calibration, given the importance of reliable predicted probabilities in clinical decision-making [42, 43]. Consequently, calibration contributes approximately 33.33% of the overall CMES, while each of the remaining dimensions contributes approximately 22.22%. The denominator of 4.5 represents the sum of the dimension weights and normalizes the final score to the interval [0, 1], where higher values indicate better overall model performance.

The composite formulation was designed to provide an interpretable summary of multiple evaluation dimensions while avoiding the dominance of any single metric within a dimension. The dimension-specific scores also ensure that fairness, discrimination, calibration, and robustness remain separately represented before their aggregation into the overall score.

#### 2.6.2 Non-parametric Statistical Analysis

The statistical comparison was conducted using the CMES as the primary endpoint. Since all six models were evaluated on the same set of ten datasets, the observations were treated as paired measurements, with each dataset serving as a statistical block. This repeated-measures structure was considered in the selection of the statistical test to account for differences in dataset characteristics.

The Friedman test was employed as a non-parametric alternative to repeated-measures analysis of variance to determine whether statistically significant differences existed among the six models. For each dataset, the models were ranked according to their CMES, with rank 1 assigned to the highest score and rank 6 assigned to the lowest score. Tied scores were assigned their average rank. The average rank for each model was then calculated across all ten datasets. Thus, a lower average rank indicates better relative performance across the evaluated datasets.

The Friedman test statistic was calculated as:

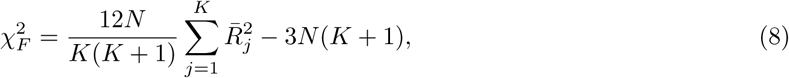

where *N* denotes the number of datasets, *K* denotes the number of models, and *R̅_j_* represents the average rank of the *j*-th model across the datasets. Under the null hypothesis, the models have equivalent performance distributions in terms of their ranks. The resulting statistic was evaluated using the chi-square approximation with *K −* 1 degrees of freedom. A significance level of *α* = 0.05 was adopted.

Kendall’s coefficient of concordance (*W*) was calculated as an effect-size measure to quantify the magnitude of agreement in the dataset-wise rankings:

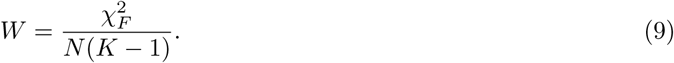

Kendall’s *W* ranges from 0 to 1, where values closer to 0 indicate weaker agreement in the rankings across datasets, while values closer to 1 indicate stronger agreement. This measure complements the Friedman test by describing the consistency of the relative model rankings.

#### 2.6.3 Post-hoc Pairwise Analysis

When the Friedman test indicated a statistically significant difference among the models (*p <* 0.05), post-hoc pairwise comparisons were conducted using the two-sided Wilcoxon signed-rank test. This test was selected because the model scores were paired by dataset and the number of datasets was relatively small. All possible pairwise comparisons among the six models were evaluated, resulting in 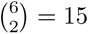 comparisons.

The Holm–Bonferroni correction was applied to the resulting *p*-values in order to account for multiple testing and control the family-wise error rate. A pairwise difference was considered statistically significant when its Holm-adjusted *p*-value was below 0.05. The pairwise analysis was performed conditionally following a significant omnibus Friedman test.

All statistical comparisons were performed using the dataset-level CMES values. Consequently, the ten datasets, rather than the individual cross-validation folds, constituted the units of comparison. This approach avoids treating fold-level measurements from the same dataset as independent observations and preserves the paired structure of the experimental design.

## 3 Results and Discussion

The Results and Discussion section presents the comparative evaluation of the models across predictive performance, fairness, calibration, and robustness, highlighting their strengths, limitations, and trade-offs across the healthcare datasets.

### 3.1 Overall Clinical Model Evaluation

The aggregated dataset-level performance of the six classifiers, summarized as mean *±* standard deviation across the ten datasets, is reported in Table 4. Across the 20 individual metrics, no single model dominated every criterion. The model-level tally of metric winners is given in Table 5. Google TabFM won eight metrics and ranked first, followed by Logistic Regression with six, TabDPT and TabPFN with two each, and TabICL and XGBoost with one each. The metrics won by TabFM mainly covered discrimination and probabilistic reliability (EOD, AUROC, Sensitivity, NPV, ECE, NLL, Brier score, and calibration slope), whereas all six metrics won by Logistic Regression related to predictive parity difference and to stability under perturbation (AUROC, accuracy, and F1 degradation, flip rate, and MAPC). The remaining fairness and calibration winners were spread across other models: TabDPT led DI and DPD, TabPFN led EqOD and calibration intercept, TabICL led PPV, and XGBoost led Specificity.

**Table 4.**
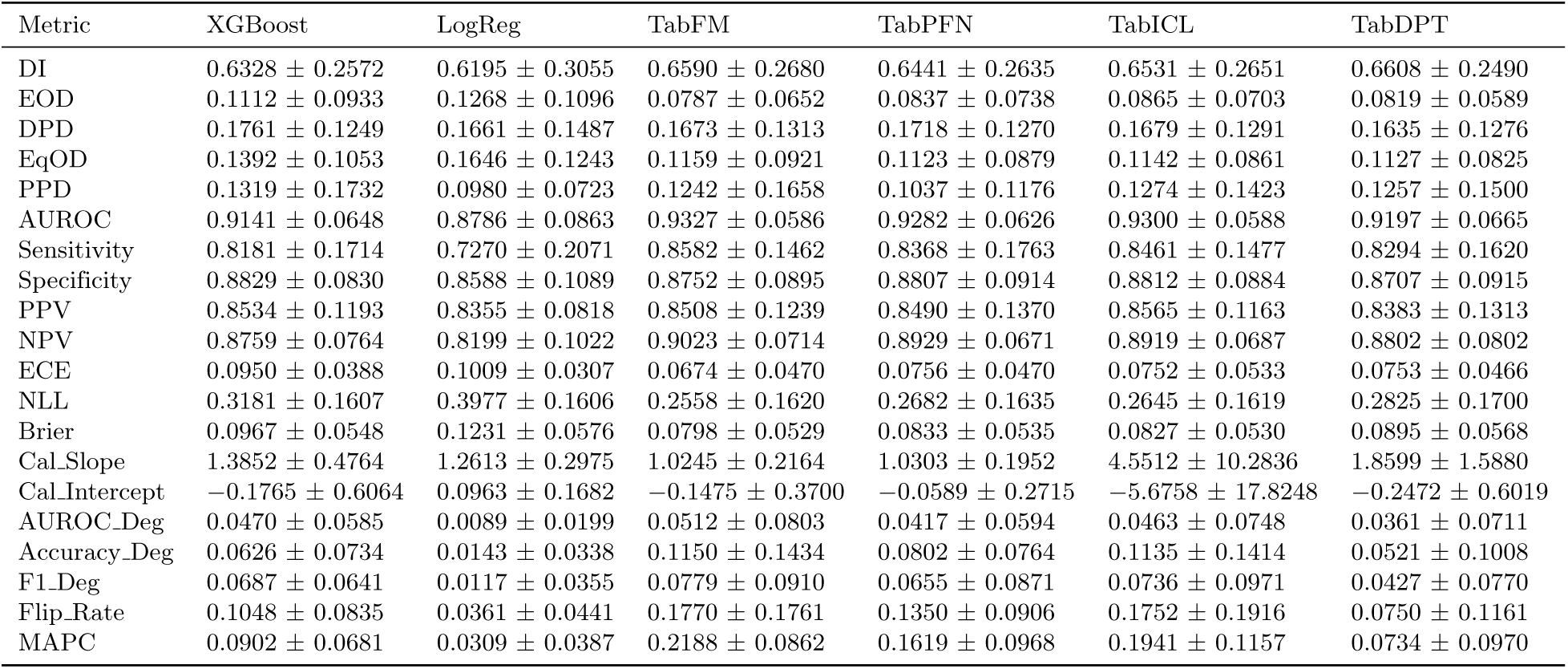
Aggregated performance of the six classification models (mean *±* SD across the ten datasets).

| Metric | XGBoost | LogReg | TabFM | TabPFN | TabICL | TabDPT |
|---|---|---|---|---|---|---|
| DI | 0.6328 ± 0.2572 | 0.6195 ± 0.3055 | 0.6590 ± 0.2680 | 0.6441 ± 0.2635 | 0.6531 ± 0.2651 | 0.6608 ± 0.2490 |
| EOD | 0.1112 ± 0.0933 | 0.1268 ± 0.1096 | 0.0787 ± 0.0652 | 0.0837 ± 0.0738 | 0.0865 ± 0.0703 | 0.0819 ± 0.0589 |
| DPD | 0.1761 ± 0.1249 | 0.1661 ± 0.1487 | 0.1673 ± 0.1313 | 0.1718 ± 0.1270 | 0.1679 ± 0.1291 | 0.1635 ± 0.1276 |
| EqOD | 0.1392 ± 0.1053 | 0.1646 ± 0.1243 | 0.1159 ± 0.0921 | 0.1123 ± 0.0879 | 0.1142 ± 0.0861 | 0.1127 ± 0.0825 |
| PPD | 0.1319 ± 0.1732 | 0.0980 ± 0.0723 | 0.1242 ± 0.1658 | 0.1037 ± 0.1176 | 0.1274 ± 0.1423 | 0.1257 ± 0.1500 |
| AUROC | 0.9141 ± 0.0648 | 0.8786 ± 0.0863 | 0.9327 ± 0.0586 | 0.9282 ± 0.0626 | 0.9300 ± 0.0588 | 0.9197 ± 0.0665 |
| Sensitivity | 0.8181 ± 0.1714 | 0.7270 ± 0.2071 | 0.8582 ± 0.1462 | 0.8368 ± 0.1763 | 0.8461 ± 0.1477 | 0.8294 ± 0.1620 |
| Specificity | 0.8829 ± 0.0830 | 0.8588 ± 0.1089 | 0.8752 ± 0.0895 | 0.8807 ± 0.0914 | 0.8812 ± 0.0884 | 0.8707 ± 0.0915 |
| PPV | 0.8534 ± 0.1193 | 0.8355 ± 0.0818 | 0.8508 ± 0.1239 | 0.8490 ± 0.1370 | 0.8565 ± 0.1163 | 0.8383 ± 0.1313 |
| NPV | 0.8759 ± 0.0764 | 0.8199 ± 0.1022 | 0.9023 ± 0.0714 | 0.8929 ± 0.0671 | 0.8919 ± 0.0687 | 0.8802 ± 0.0802 |
| ECE | 0.0950 ± 0.0388 | 0.1009 ± 0.0307 | 0.0674 ± 0.0470 | 0.0756 ± 0.0470 | 0.0752 ± 0.0533 | 0.0753 ± 0.0466 |
| NLL | 0.3181 ± 0.1607 | 0.3977 ± 0.1606 | 0.2558 ± 0.1620 | 0.2682 ± 0.1635 | 0.2645 ± 0.1619 | 0.2825 ± 0.1700 |
| Brier | 0.0967 ± 0.0548 | 0.1231 ± 0.0576 | 0.0798 ± 0.0529 | 0.0833 ± 0.0535 | 0.0827 ± 0.0530 | 0.0895 ± 0.0568 |
| Cal_Slope | 1.3852 ± 0.4764 | 1.2613 ± 0.2975 | 1.0245 ± 0.2164 | 1.0303 ± 0.1952 | 4.5512 ± 10.2836 | 1.8599 ± 1.5880 |
| Cal_Intercept | −0.1765 ± 0.6064 | 0.0963 ± 0.1682 | −0.1475 ± 0.3700 | −0.0589 ± 0.2715 | −5.6758 ± 17.8248 | −0.2472 ± 0.6019 |
| AUROC Deg | 0.0470 ± 0.0585 | 0.0089 ± 0.0199 | 0.0512 ± 0.0803 | 0.0417 ± 0.0594 | 0.0463 ± 0.0748 | 0.0361 ± 0.0711 |
| Accuracy Deg | 0.0626 ± 0.0734 | 0.0143 ± 0.0338 | 0.1150 ± 0.1434 | 0.0802 ± 0.0764 | 0.1135 ± 0.1414 | 0.0521 ± 0.1008 |
| F1 Deg | 0.0687 ± 0.0641 | 0.0117 ± 0.0355 | 0.0779 ± 0.0910 | 0.0655 ± 0.0871 | 0.0736 ± 0.0971 | 0.0427 ± 0.0770 |
| Flip Rate | 0.1048 ± 0.0835 | 0.0361 ± 0.0441 | 0.1770 ± 0.1761 | 0.1350 ± 0.0906 | 0.1752 ± 0.1916 | 0.0750 ± 0.1161 |
| MAPC | 0.0902 ± 0.0681 | 0.0309 ± 0.0387 | 0.2188 ± 0.0862 | 0.1619 ± 0.0968 | 0.1941 ± 0.1157 | 0.0734 ± 0.0970 |

**Table 5.** Model ranking based on the number of individual metrics for which each model achieved the best mean value.

| Rank | Model | Metrics won | Won metrics |
|---|---|---|---|
| 1 | TabFM | 8 | EOD, AUROC, Sensitivity, NPV, ECE, NLL, Brier, Cal_Slope |
| 2 | LogReg | 6 | PPD, AUROC_Deg, Accuracy_Deg, F1_Deg, Flip_Rate, MAPC |
| 3 | TabDPT | 2 | DI, DPD |
| 3 | TabPFN | 2 | EqOD, Cal_Intercept |
| 5 | TabICL | 1 | PPV |
| 5 | XGBoost | 1 | Specificity |

Figure 1 (A) presents the dataset-level CMES for every model–dataset combination, together with the mean across datasets. The mean CMES was highest for TabFM (0.881), followed closely by TabDPT (0.880), TabICL (0.877), TabPFN (0.876), XGBoost (0.865), and Logistic Regression (0.855). Scores were highest in neurology and endocrinology, particularly for Autism, Diabetes, and Alzheimer’s disease (0.90–0.96), and lowest in Hepatitis (0.74–0.78) and in Heart (0.79–0.81). The best model per dataset varied: TabFM led in Stroke, Alzheimer’s disease, COPD, Diabetes, and Liver. TabDPT led in Heart, Thyroid, and Hepatitis. TabPFN led in Arrhythmia, and TabICL led in Autism. The largest between-model spread occurred in Arrhythmia, where Logistic Regression (0.78) trailed TabPFN (0.85) by a considerable margin, and in Hepatitis, where TabPFN obtained 0.74 compared with 0.78 for TabDPT. Figure 1 (B) quantifies the paired CMES improvement of each model over the Logistic Regression baseline. TabFM achieved the largest mean gain (+0.026), followed by TabDPT (+0.025), TabICL (+0.023), TabPFN (+0.021), and XGBoost (+0.010). The bootstrap intervals of the four foundation models lay clearly above zero, whereas the interval of XGBoost reached approximately zero at its lower bound, which suggests that the advantage of XGBoost over Logistic Regression is the least certain of the five. Figure 1 (C) illustrates how often each model occupied each rank within a dataset. TabFM ranked first in five datasets and TabDPT in three, whereas TabICL and TabPFN were first in one dataset each. Logistic Regression ranked last in seven datasets, XGBoost was fifth in five datasets, and TabPFN ranked last in two. The resulting mean ranks were 2.0 for TabFM, 2.4 for TabDPT, 2.9 for TabICL, 3.8 for TabPFN, 4.4 for XGBoost, and 5.5 for Logistic Regression, which separates the foundation models from the two conventional baselines and places TabFM consistently near the top.

**Fig 1.**
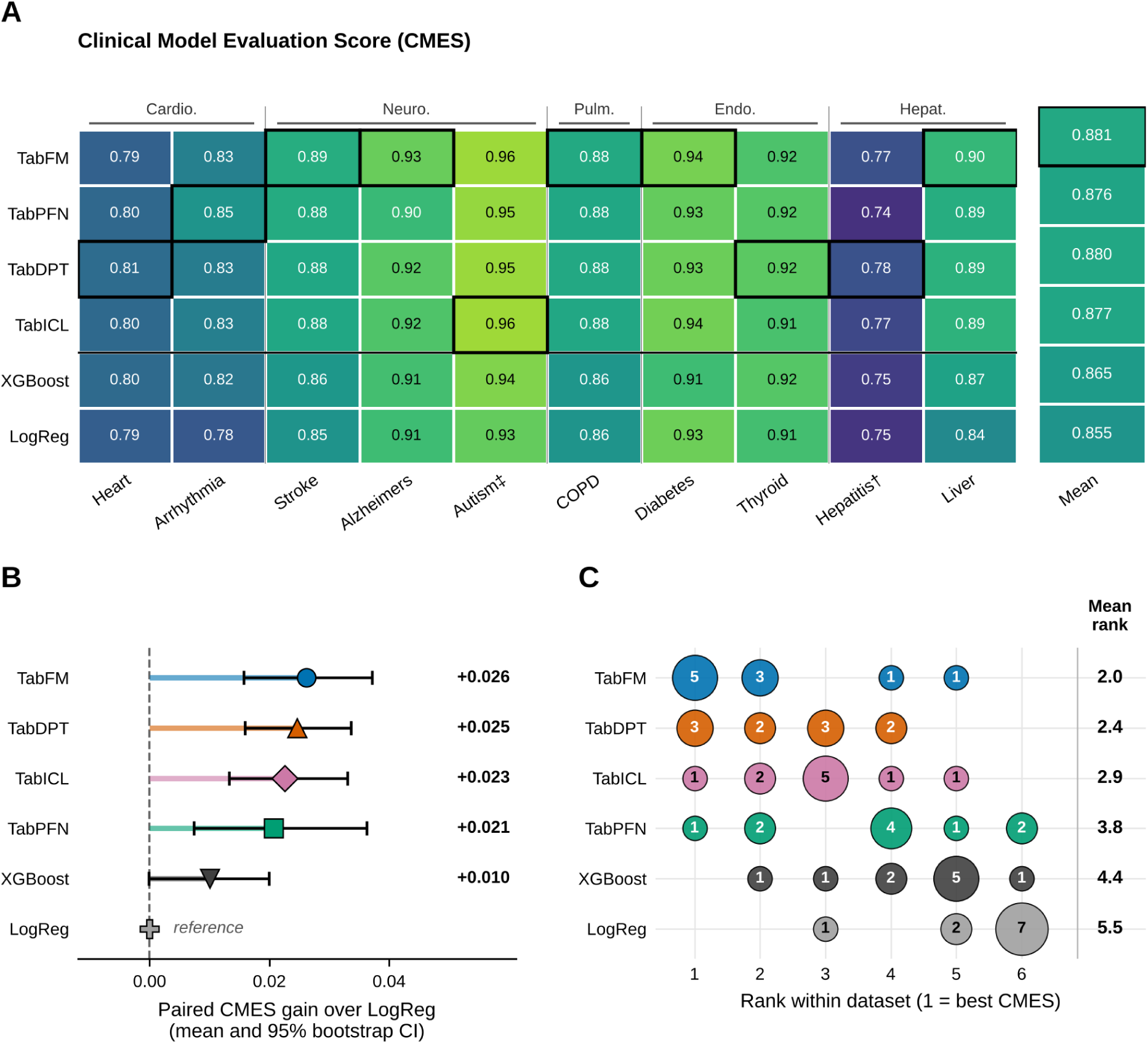
Overall CMES. (A) Dataset-level CMES heatmap grouped by clinical domain, with black boxes marking the best model per dataset. (B) Paired CMES gain over Logistic Regression with 95% bootstrap confidence intervals. (C) Distribution of within-dataset ranks and mean rank per model.

### 3.2 Trade-offs Across Evaluation Dimensions

Figure 2 (A) displays the mean composite score of each model for fairness, predictive performance, calibration, and robustness. The dimensions differed markedly in their ranges. All models scored within a narrow band on fairness (approximately 0.73–0.75, with TabDPT highest and Logistic Regression lowest). The spread was largest for predictive performance, where TabFM (approximately 0.90) and the other foundation models (0.88–0.89) clearly exceeded XGBoost (approximately 0.87) and Logistic Regression (approximately 0.80). For calibration, TabFM was highest (approximately 0.93), the remaining foundation models were around 0.92, and XGBoost and Logistic Regression obtained approximately 0.90 and 0.88. The ordering reversed for robustness, where Logistic Regression obtained by far the highest score (approximately 0.99), followed by TabDPT (approximately 0.96), while TabFM had the lowest value (approximately 0.93).

**Fig 2.**
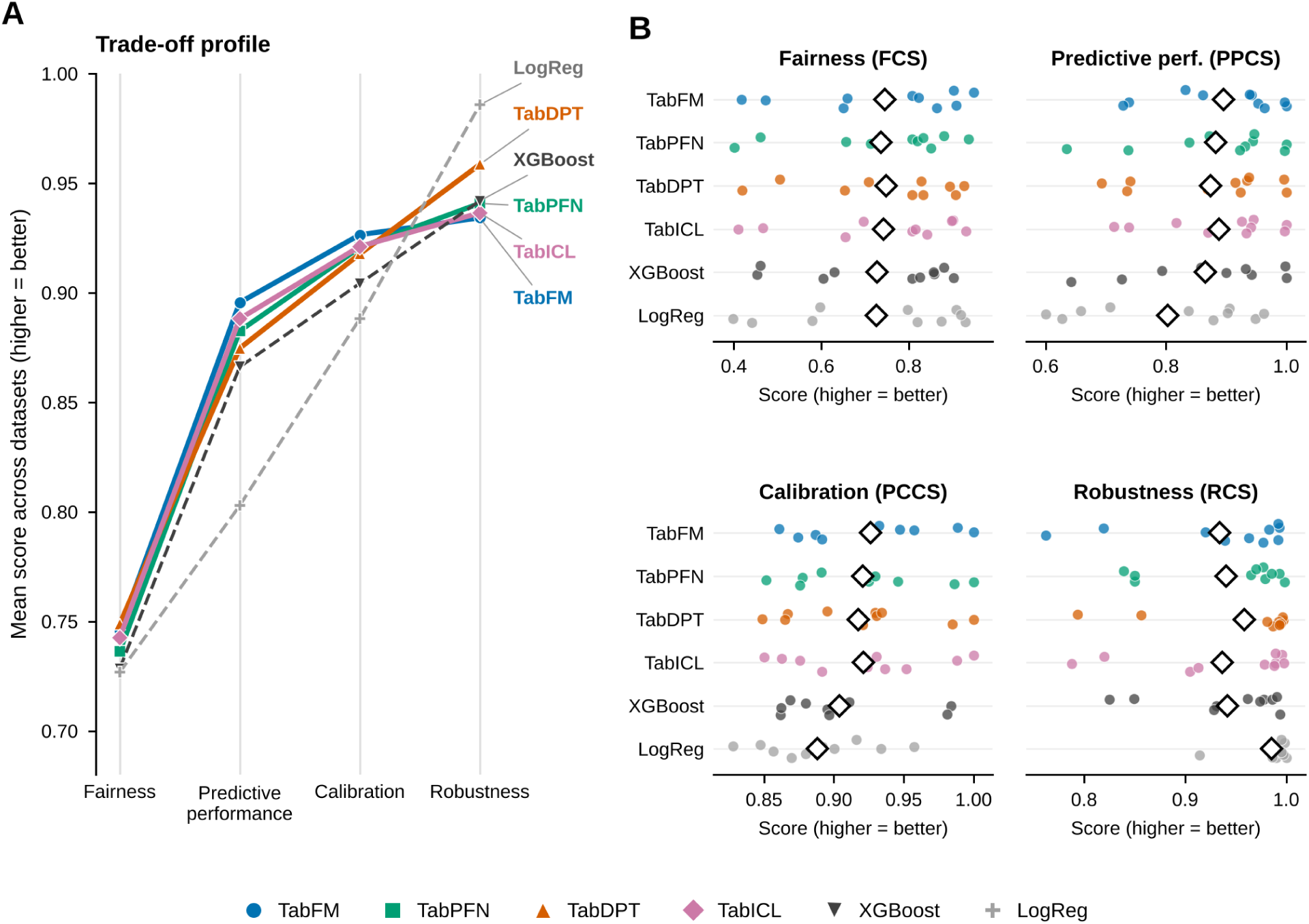
Dimension-specific composite scores. (A) Trade-off profile showing the mean score of each model across the four dimensions. (B) Dataset-level distributions of FCS, PPCS, PCCS, and RCS, with diamonds marking the mean across the ten datasets.

Figure 2 (B) visualizes the dataset-level spread behind these means. Fairness showed the widest dispersion, with individual dataset scores ranging from approximately 0.40 to 0.95 for every model, so the differences between model means were small compared with the variability across datasets. Predictive performance was concentrated near 0.85–1.0 for the foundation models, with a few lower values for TabPFN and for the baselines (approximately 0.60–0.75). Calibration scores were generally confined to between 0.83 and 1.0, and robustness scores were concentrated close to 1.0 with several low outliers for the foundation models, notably TabFM, TabICL, and TabDPT (approximately 0.76–0.86).

### 3.3 Discrimination

Figure 3 (A) reports the AUROC of each model for every dataset. In most datasets the foundation models were close to one another and above XGBoost and Logistic Regression. The clearest separation occurred in Liver, where the foundation models reached an AUROC of approximately 0.97, XGBoost approximately 0.91, and Logistic Regression only approximately 0.71, and in Arrhythmia, TabFM (approximately 0.91) and TabPFN (approximately 0.90) exceeded XGBoost (approximately 0.87) and Logistic Regression (approximately 0.79), while TabDPT was visibly weaker among the foundation models (approximately 0.82). Autism was effectively saturated for all models (AUROC *≈* 1.0), Stroke showed overlapping and wide intervals for all models (approximately 0.78–0.86), and Hepatitis exhibited wide fold-to-fold intervals that overlapped across models, in line with its small sample size. Averaged over datasets (Table 4), TabFM obtained the highest AUROC (0.9327), followed by TabICL (0.9300), TabPFN (0.9282), TabDPT (0.9197), XGBoost (0.9141), and Logistic Regression (0.8786).

**Fig 3.**
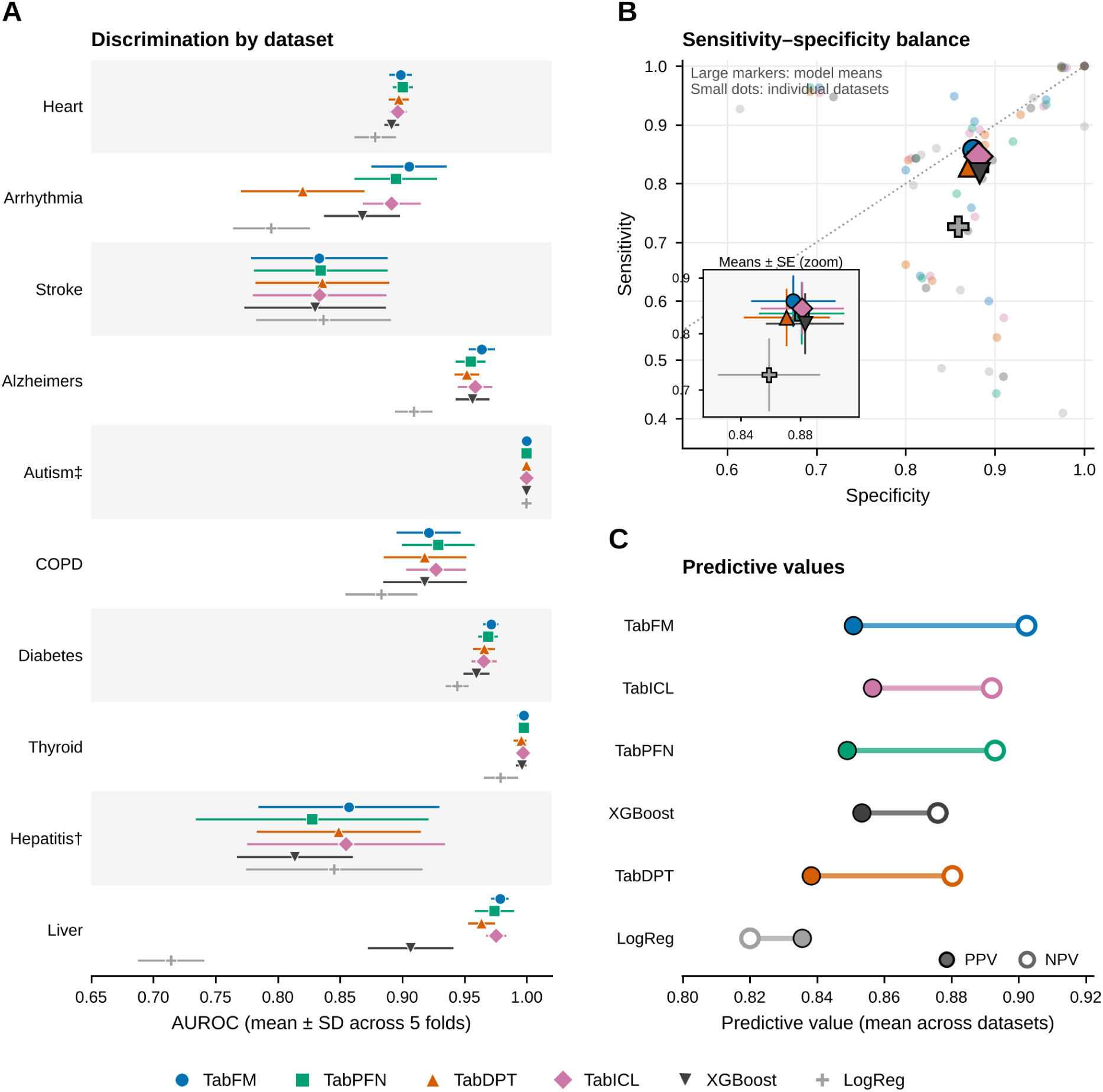
Discrimination performance. (A) AUROC by dataset (mean *±* SD across five folds). (B) Sensitivity–specificity balance, with large markers showing model means and an inset showing means *±* SE. (C) Positive and negative predictive values averaged across datasets.

Figure 3 (B) visualizes the balance between sensitivity and specificity. The foundation AI models and XGBoost formed a tight cluster at a specificity of approximately 0.87–0.88 and a sensitivity of 0.82–0.86, with TabFM reaching the highest mean sensitivity (0.8582) and XGBoost the highest specificity (0.8829). Logistic Regression was clearly separated from this cluster, with a markedly lower sensitivity (0.7270) than specificity (0.8588), which places it well below the diagonal of equal sensitivity and specificity and indicates a tendency to miss positive cases. The zoomed inset confirms that the standard-error bars of the foundation models overlap, whereas the Logistic Regression interval is well separated. The small dots further show that individual datasets varied considerably around these means, with some datasets near perfect classification and others with sensitivity below 0.6.

Figure 3 (C) depicts the positive and negative predictive values of each model. For all models except Logistic Regression, the NPV exceeded the PPV, and the gap was largest for TabFM (NPV 0.9023 versus PPV 0.8508), followed by TabPFN (0.8929 versus 0.8490) and TabICL (0.8919 versus 0.8565). The PPV values of the foundation models and XGBoost were similar (0.8383–0.8565). Logistic Regression was the only model with a PPV (0.8355) higher than its NPV (0.8199), and its NPV was the lowest of all models, meaning that negative predictions from this baseline were the least reliable.

### 3.4 Calibration and Probabilistic Accuracy

Figure 4 (A) summarizes the calibration error and probabilistic accuracy at the dataset level. Hepatitis produced the largest bubbles for every model, reflecting the instability of probability estimates when fewer test cases are available per fold. In contrast, the ECE was nearly zero for the foundation models in Autism, and the bubbles for Thyroid were small and pale for all models except Logistic Regression. Logistic Regression had the darkest bubbles (highest Brier scores) in Arrhythmia and Liver, which coincides with its weak discrimination in those datasets, and XGBoost showed larger errors than the foundation models in Alzheimer’s disease and Diabetes. Overall, the foundation models displayed smaller bubbles and lighter colours than the baselines in most datasets, which is consistent with their lower mean ECE (0.0674–0.0756 versus 0.0950 for XGBoost and 0.1009 for Logistic Regression) and Brier score (0.0798–0.0895 versus 0.0967 and 0.1231) as shown in Table 4. Figure 4 (B) compares the distribution of NLL across datasets. TabFM had the lowest mean (0.2558), followed by TabICL (0.2645), TabPFN (0.2682), and TabDPT (0.2825), and the medians of the first three foundation models were nearly identical (approximately 0.25). XGBoost (0.3181) and Logistic Regression (0.3977) were visibly higher, and Logistic Regression additionally exhibited the highest median, the longest upper whisker, and the largest dispersion, indicating that its probabilistic accuracy deteriorated sharply in particular datasets. Figure 4 (C) plots the calibration slope against the calibration intercept, with the green zone indicating acceptable calibration (slope between 0.8 and 1.25, intercept within *±*0.2). Most points of all models were concentrated in or near this zone, and TabFM and TabPFN had mean slopes closest to the ideal value (1.0245 and 1.0303, respectively). A few extreme cases were located far from the zone, namely TabDPT in Thyroid and Autism, and TabICL in Autism, and one point was outside the plotted range (TabICL in Thyroid, intercept *−*56.4, slope 33.7). These extreme values inflate the mean slope and intercept of TabICL (4.5512 and *−*5.6758) and, to a lesser extent, of TabDPT (1.8599 and *−*0.2472) in Table 4.

**Fig 4.**
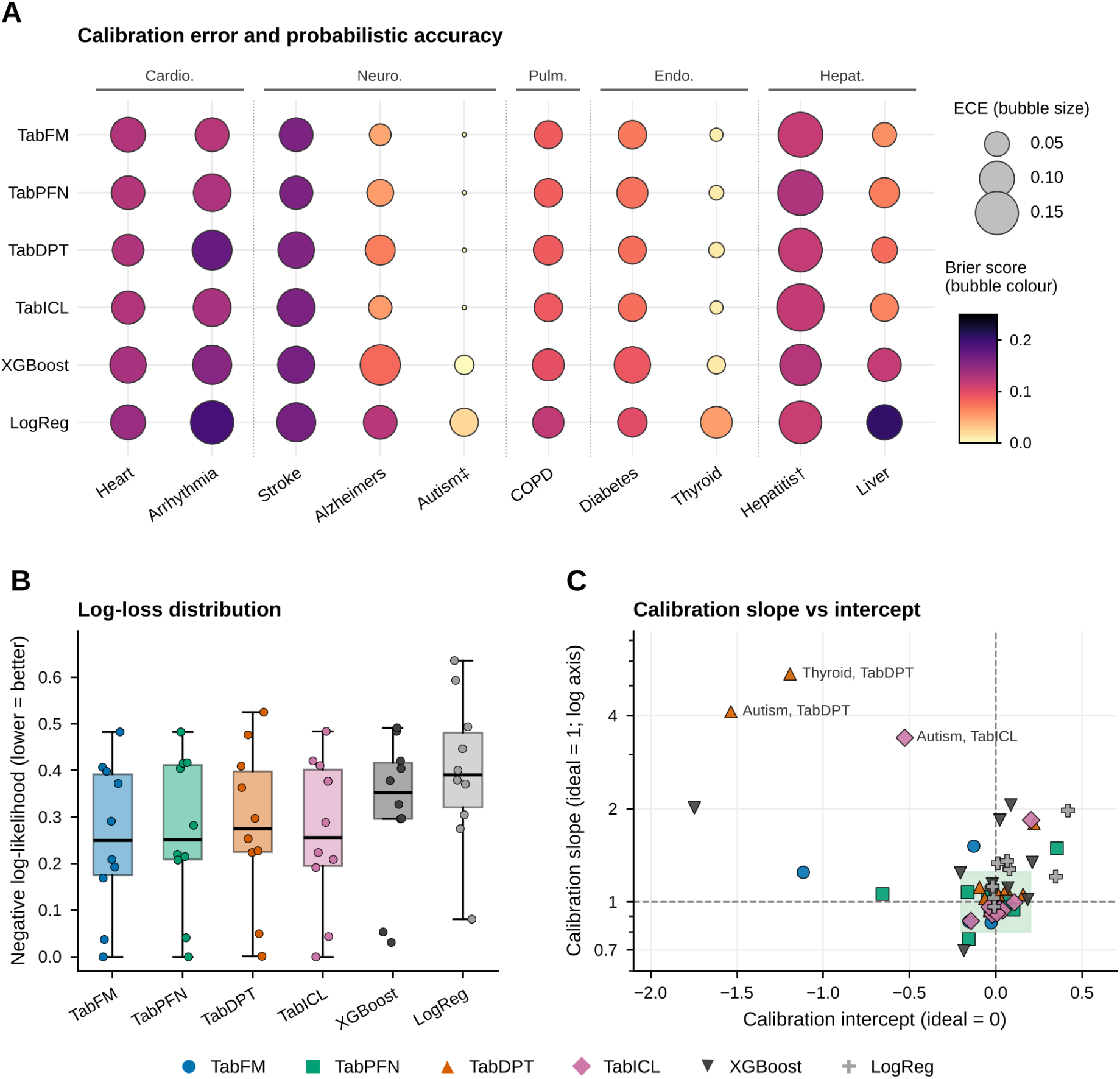
Calibration and probabilistic accuracy. (A) ECE (bubble size) and Brier score (bubble colour) for each model and dataset. (B) Distribution of NLL across datasets. (C) Calibration slope versus intercept.

### 3.5 Fairness

Figure 5 (A) shows the DI of each model by dataset against the conventional 0.80 threshold. The datasets fell into three groups. Heart, Arrhythmia, COPD, Autism, and Hepatitis were below the threshold for essentially all models, with DI ranging from approximately 0.25–0.32 in Heart, 0.44–0.63 in Arrhythmia, 0.57–0.64 in COPD, and 0.64–0.71 in Autism. Hepatitis was the most extreme case, with DI between zero (Logistic Regression) and approximately 0.17 (TabDPT). Stroke, Diabetes, and Liver were above the threshold for most models, while Alzheimer’s disease and Thyroid were close to the boundary (approximately 0.75–0.85 and 0.75–0.78, respectively). This pattern suggests that disparity originates largely from the data and the clinical task rather than from a particular algorithm. Figure 5 (B) summarizes the mean disparity across the five fairness metrics, where lower values are fairer. TabDPT obtained the best values for 1 *–* DI (0.34) and DPD (0.16), TabFM was best for EOD (0.08), TabPFN was best for EqOD (0.11), and Logistic Regression was best for PPD (0.10). The differences between the foundation models were very small for all metrics, with 1 *−* DI ranging from 0.34 to 0.36 and DPD from 0.16 to 0.17. The baselines were mostly worse on the equalized-odds-type metrics, with XGBoost and Logistic Regression obtaining the highest values for EOD (0.11 and 0.13) and EqOD (0.14 and 0.16), and Logistic Regression was at the same time the least fair model on 1 *−* DI (0.38), which is consistent with the small differences in the fairness composite shown in Figure 2. Figure 5 (C) relates DI to discrimination. Points of the same dataset formed vertical bands, which shows that DI was determined mainly by the dataset rather than by the model’s AUROC. Autism, where the AUROC was equal to 1, yielded DI values of approximately 0.65–0.78 and therefore remained below or close to the threshold, whereas Heart combined an AUROC of approximately 0.90 with DI of only 0.25–0.32, and Hepatitis produced DI of 0–0.17 at AUROC values of approximately 0.82–0.86. In the upper part of the plot, the Logistic Regression point in Liver had a high DI (approximately 0.88) despite a low AUROC (approximately 0.71). Taken together, these patterns indicate that improved predictive performance alone does not resolve DI.

**Fig 5.**
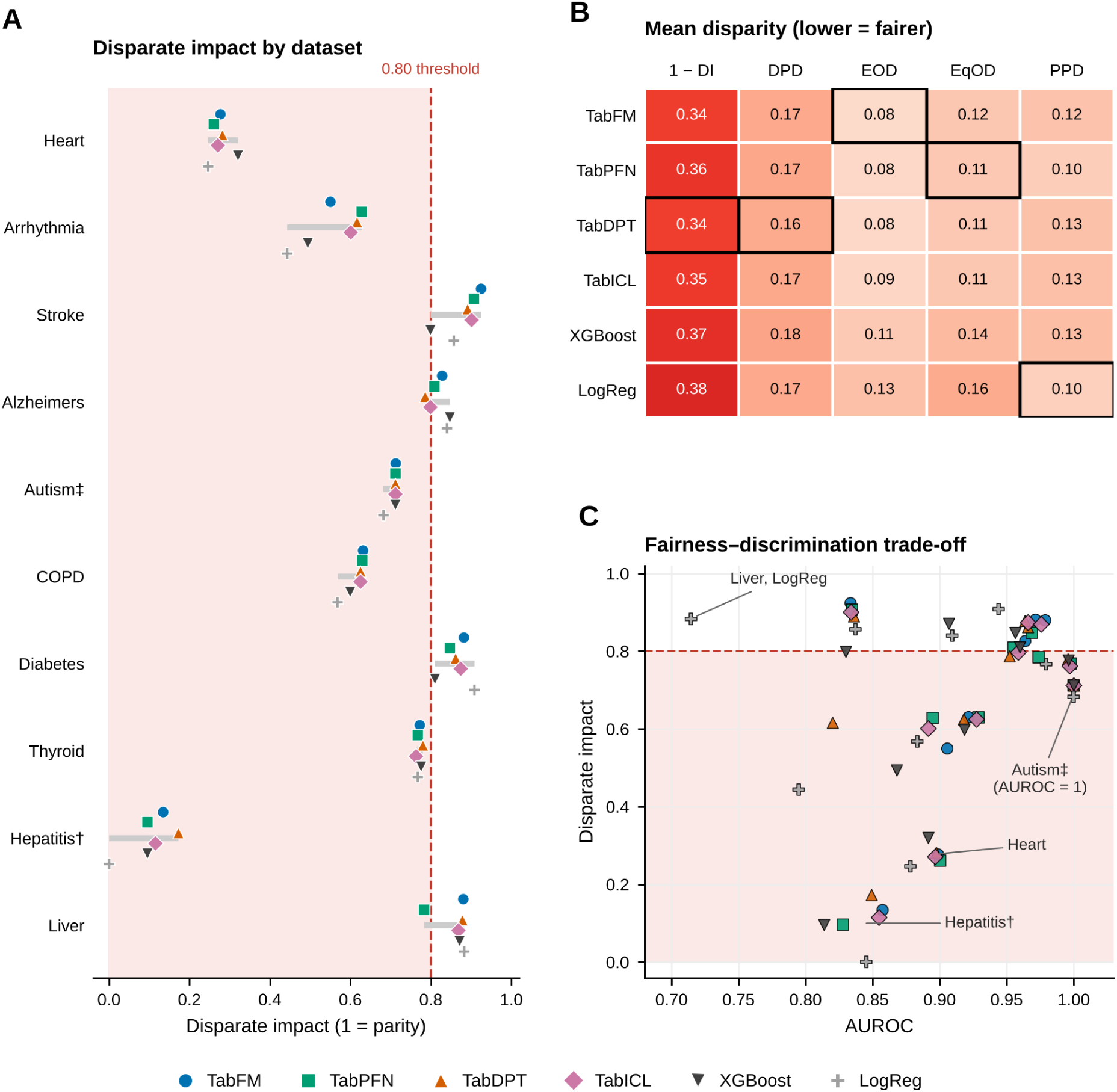
Fairness assessment. (A) DI by dataset relative to the 0.80 threshold. (B) Mean disparity metrics for each model, with black boxes marking the best model per metric. (C) Fairness–discrimination trade-off, plotting DI against AUROC.

### 3.6 Robustness to Input Perturbation

Figure 6 (A) illustrates the loss of discrimination when Gaussian noise equal to 10% of the training-set feature standard deviation was added to the test features. On clean data the foundation AI models achieved a mean AUROC of approximately 0.92–0.93 and XGBoost approximately 0.91, compared with approximately 0.88 for Logistic Regression. Under noise, the AUROC decreased by 0.051 for TabFM, 0.047 for XGBoost, 0.046 for TabICL, 0.042 for TabPFN, 0.036 for TabDPT, and only 0.009 for Logistic Regression. As a result, the noisy-data AUROC of the foundation models (approximately 0.88–0.89) was only marginally above that of Logistic Regression (approximately 0.87), and XGBoost fell slightly below Logistic Regression, so the advantage of the foundation models in discrimination was substantially reduced, but not removed, by noise. Figure 6 (B) visualizes prediction instability by relating the flip rate to the mean absolute change in predicted probability. Logistic Regression and TabDPT points were concentrated near the origin in most datasets, in agreement with the low mean flip rates (0.0361 and 0.0750) and MAPC values (0.0309 and 0.0734) of these models. The other foundation models and XGBoost spread out along an increasing relationship, with the highest instability for TabFM and TabICL in Arrhythmia (flip rates of approximately 0.55–0.60), for TabFM in Liver, and for TabICL in Thyroid. TabFM had the largest mean MAPC (0.2188) and, together with TabICL, the largest mean flip rate (0.1770 and 0.1752, respectively). Figure 6 (C) breaks down the AUROC degradation by dataset. The loss was highly localized: for most datasets and models the degradation was between 0.00 and 0.04, but Thyroid and Liver showed substantially larger values for the foundation models and XGBoost, reaching 0.16 and 0.24 for TabFM, 0.15 and 0.22 for TabICL, 0.12 and 0.21 for TabDPT, 0.12 and 0.16 for TabPFN, and 0.12 and 0.18 for XGBoost. Logistic Regression was largely unaffected, with only a moderate loss in Thyroid (0.06). An additional isolated loss was seen for TabPFN in Alzheimer’s disease (0.09). The mean degradation was 0.05 for TabFM, TabICL, and XGBoost, 0.04 for TabPFN and TabDPT, and only 0.01 for Logistic Regression.

**Fig 6.**
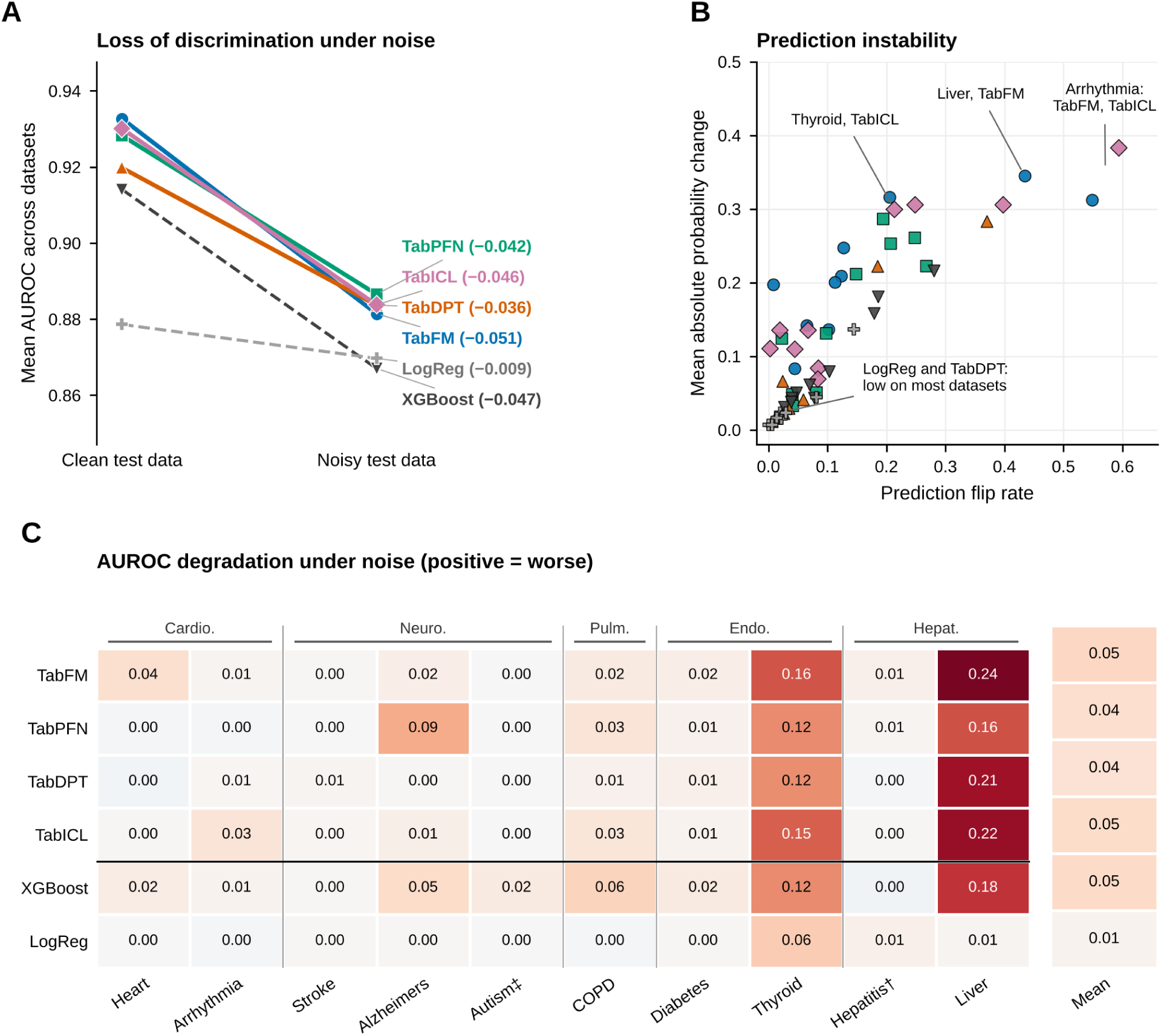
Robustness to feature noise. (A) Mean AUROC on clean and noisy test data. (B) Prediction instability, expressed as flip rate versus mean absolute probability change. (C) AUROC degradation under noise by model and dataset.

### 3.7 Statistical Comparison of Models

The omnibus Friedman test on the dataset-level CMES is reported in Table 6. The test detected a statistically significant difference among the six models 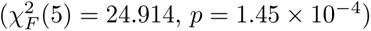, and Kendall’s *W* = 0.498 indicates a moderate degree of agreement between the datasets in how the models were ranked.

**Table 6.**
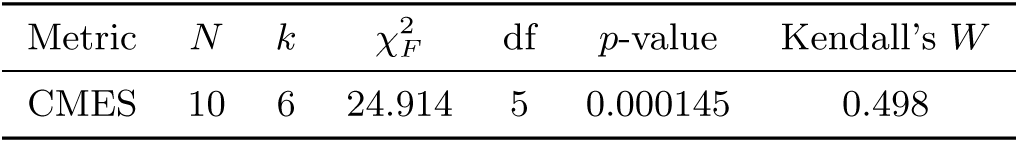
Friedman test for differences in CMES among the six models.

The mean CMES, its standard deviation, and the mean Friedman rank of every model are listed in Table 7. The ordering by mean CMES and by mean rank was identical: TabFM (0.8811; rank 2.0) was followed by TabDPT (0.8796; 2.4), TabICL (0.8775; 2.9), TabPFN (0.8756; 3.8), XGBoost (0.8650; 4.4), and Logistic Regression (0.8549; 5.5). The standard deviation of CMES across datasets was similar for all models (0.059–0.066), which means that the differences in mean CMES are small relative to the variation between datasets.

**Table 7.** Mean CMES, standard deviation, and mean Friedman rank of the six models (lower rank is better).

| Model | Mean CMES | SD CMES | Mean rank |
|---|---|---|---|
| TabFM | 0.8811 | 0.0620 | 2.0 |
| TabDPT | 0.8796 | 0.0593 | 2.4 |
| TabICL | 0.8775 | 0.0611 | 2.9 |
| TabPFN | 0.8756 | 0.0627 | 3.8 |
| XGBoost | 0.8650 | 0.0591 | 4.4 |
| Logistic Regression | 0.8549 | 0.0659 | 5.5 |

The post-hoc pairwise comparisons are presented in Table 8. After Holm adjustment, four of the 15 comparisons remained significant. Logistic Regression was significantly worse than TabFM (mean difference *−*0.026, adjusted *p* = 0.029), TabICL (*−*0.023, adjusted *p* = 0.029), and TabDPT (*−*0.025, adjusted *p* = 0.047), and TabDPT was significantly better than XGBoost (+0.015, adjusted *p* = 0.029). The comparisons of TabFM with XGBoost (adjusted *p* = 0.107) and TabICL with XGBoost (adjusted *p* = 0.137) were nominally significant before correction but did not survive it, Logistic Regression versus TabPFN (adjusted *p* = 0.246) and versus XGBoost (adjusted *p* = 0.391) were not significant, and no pair of foundation models differed significantly.

**Table 8.**
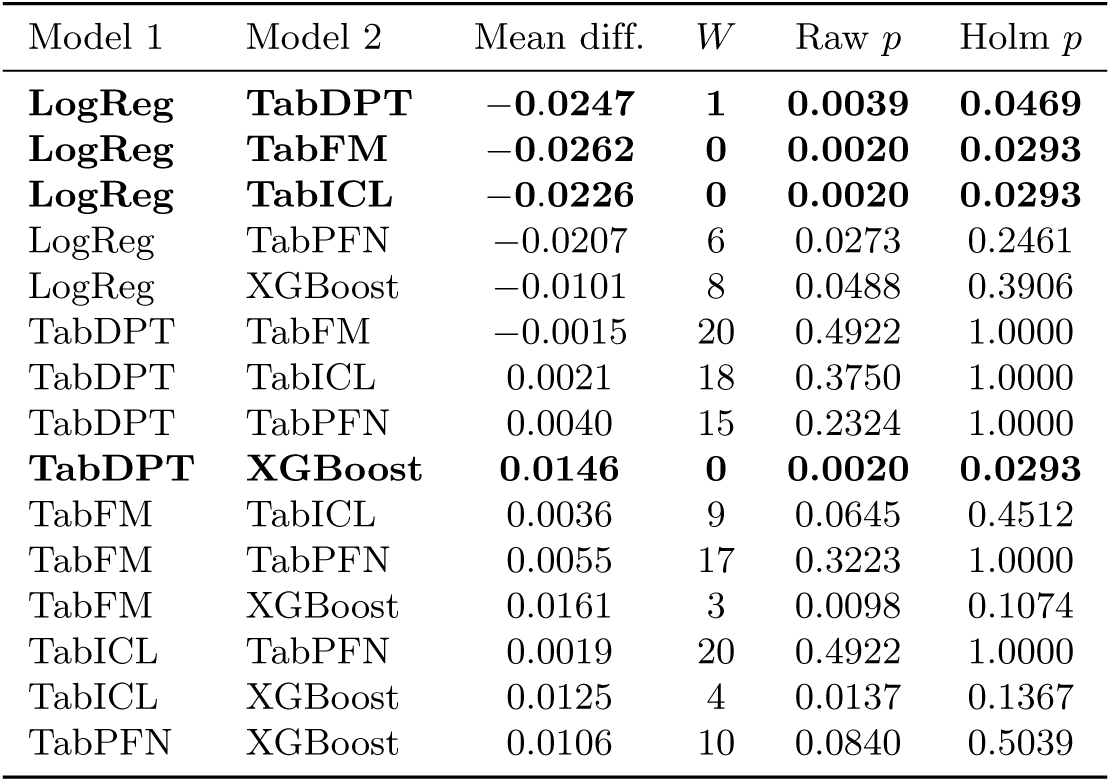
Post-hoc pairwise Wilcoxon signed-rank comparisons of CMES with Holm adjustment. The mean difference is Model 1 minus Model 2. Significant results (adjusted *p <* 0.05) are in bold.

| Model 1 | Model 2 | Mean diff. | *W* | Raw *p* | Holm *p* |
|---|---|---|---|---|---|
| **LogReg** | **TabDPT** | **−0.0247** | **1** | **0.0039** | **0.0469** |
| **LogReg** | **TabFM** | **−0.0262** | **0** | **0.0020** | **0.0293** |
| **LogReg** | **TabICL** | **−0.0226** | **0** | **0.0020** | **0.0293** |
| LogReg | TabPFN | −0.0207 | 6 | 0.0273 | 0.2461 |
| LogReg | XGBoost | −0.0101 | 8 | 0.0488 | 0.3906 |
| TabDPT | TabFM | −0.0015 | 20 | 0.4922 | 1.0000 |
| TabDPT | TabICL | 0.0021 | 18 | 0.3750 | 1.0000 |
| TabDPT | TabPFN | 0.0040 | 15 | 0.2324 | 1.0000 |
| **TabDPT** | **XGBoost** | **0.0146** | **0** | **0.0020** | **0.0293** |
| TabFM | TabICL | 0.0036 | 9 | 0.0645 | 0.4512 |
| TabFM | TabPFN | 0.0055 | 17 | 0.3223 | 1.0000 |
| TabFM | XGBoost | 0.0161 | 3 | 0.0098 | 0.1074 |
| TabICL | TabPFN | 0.0019 | 20 | 0.4922 | 1.0000 |
| TabICL | XGBoost | 0.0125 | 4 | 0.0137 | 0.1367 |
| TabPFN | XGBoost | 0.0106 | 10 | 0.0840 | 0.5039 |

Figure 7 (A) visualizes the mean ranks on a common axis together with the Friedman test result. TabFM, TabDPT, and TabICL were located at the favourable end (2.0–2.9), TabPFN was positioned between these models and the baselines (3.8), and XGBoost and Logistic Regression were separated at 4.4 and 5.5. Three overlapping bars join the models whose CMES does not differ significantly: the first bar links TabFM, TabDPT, TabICL, and TabPFN, the second links TabICL, TabPFN, and XGBoost, and the third links TabPFN, XGBoost, and Logistic Regression. The significance bars represent selected maximal nonsignificant groups rather than all pairwise nonsignificant comparisons. Figure 7 (B) depicts the Holm-adjusted *p*-values as a lower-triangular matrix. All comparisons involving TabPFN with TabFM, TabDPT, and TabICL yielded adjusted *p*-values of 1.00, as did TabDPT versus TabICL. The four significant cells (marked with an asterisk) were Logistic Regression versus TabFM (0.029), TabDPT (0.047), and TabICL (0.029), and XGBoost versus TabDPT (0.029), while TabFM versus XGBoost (0.11) and TabICL versus XGBoost (0.14) did not reach significance. Figure 7 (C) traces the rank of each model across the ten datasets, grouped by clinical domain. The trajectories crossed frequently, so no foundation model was consistently best. TabFM ranked first in five datasets but fifth in Heart, TabDPT ranked first in Heart, Thyroid, and Hepatitis but fourth in Arrhythmia and Diabetes, and TabPFN ranked first in Arrhythmia but last in Alzheimer’s disease and Hepatitis. TabICL was the most stable model among the foundation models, remaining between first and fifth place. XGBoost reached second place only in Heart and was last in Diabetes, and Logistic Regression occupied the last rank in seven datasets, which accounts for its mean rank of 5.5.

**Fig 7.**
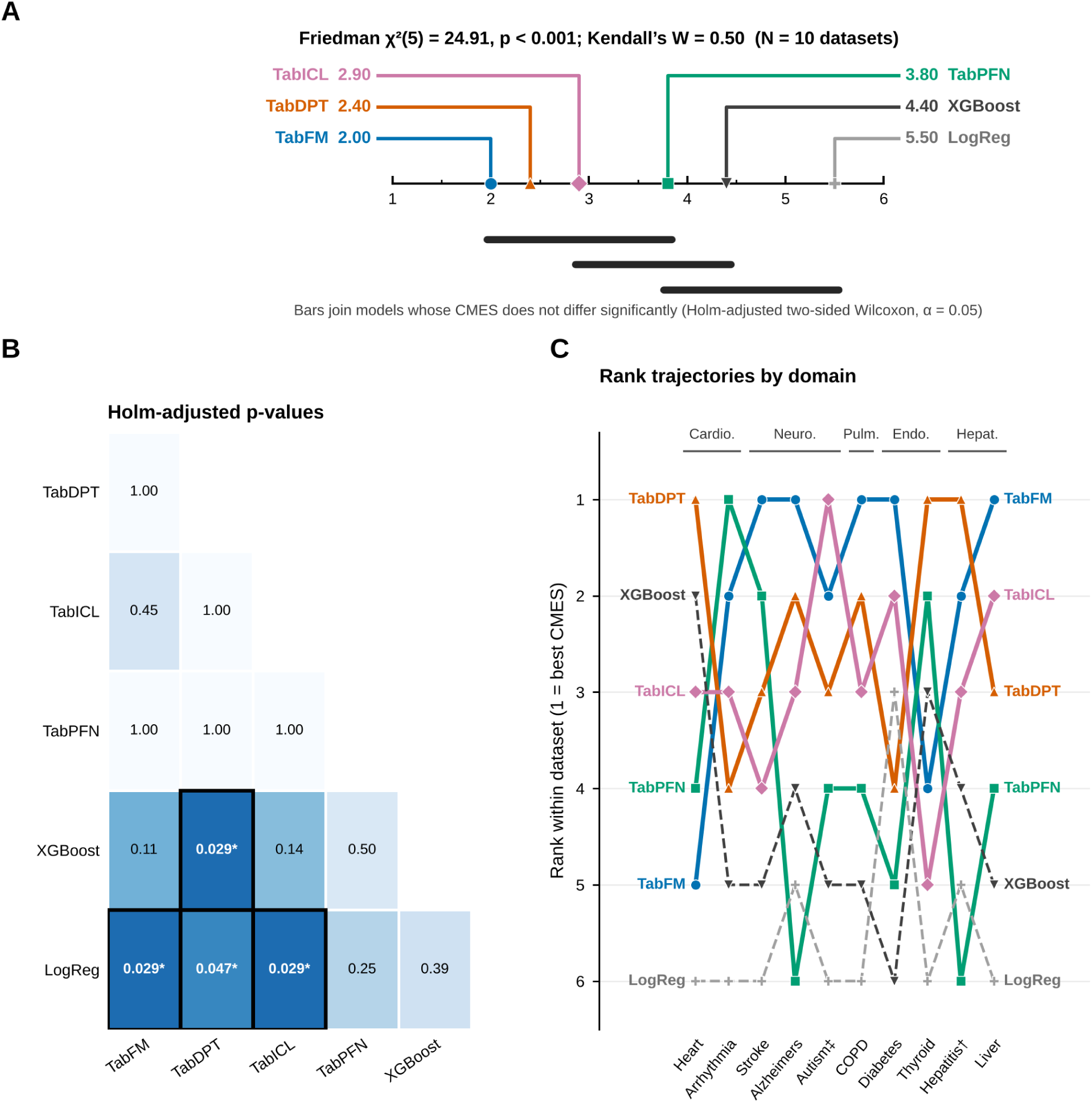
Statistical comparison of models based on dataset-level CMES. (A) Critical-difference-style diagram of mean ranks with the Friedman test summary. (B) Holm-adjusted pairwise *p*-values. (C) Rank trajectories of each model across datasets and clinical domains.

### 3.8 Discussion

The results indicate that the tabular foundation AI models offered a consistent advantage in discrimination and probabilistic reliability over the conventional baselines, but only a limited advantage in fairness and a disadvantage in robustness. Google TabFM was the strongest model overall, achieving the highest mean AUROC, sensitivity, and NPV and the lowest ECE, NLL, and Brier score (Table 5), together with the best mean CMES and rank (Table 7). The advantage was most apparent in Liver and Arrhythmia, where the AUROC of Logistic Regression was markedly lower (Figure 3 (A)), and in the sensitivity of the models, where Logistic Regression missed a substantially larger proportion of positive cases (Figure 3 (B)).

The robustness findings qualify this picture. The models with the best discrimination and calibration were also the most affected by feature noise, and the degradation was concentrated in the Thyroid and Liver datasets (Figure 6 (C)). The near-perfect robustness of Logistic Regression should be interpreted accordingly: its low flip rate and probability change (Figure 6 (B)) partly reflect a lower-capacity model that already performs worse on clean data (Figure 6 (A)), and the robustness composite rewards stability irrespective of the baseline performance level. After noise, the advantage of the foundation models in AUROC over Logistic Regression shrank to approximately 0.01–0.02, and XGBoost fell below Logistic Regression. TabDPT combined a competitive discrimination with the smallest degradation, flip rate, and MAPC among the foundation models, which suggests that stability is not an inherent trade-off of the foundation-model class.

The fairness results highlight that DI was largely dataset-driven. Five datasets (Heart, Arrhythmia, COPD, Autism, and Hepatitis) fell below the 0.80 threshold for virtually all models (Figure 5 (A)), and no model was best across all fairness metrics (Figure 5 (B)). The relationship between discrimination and DI was weak (Figure 5 (C)), and the fairness composite scores were nearly identical across models (Figure 2 (A)). These findings suggest that adopting a foundation model neither improves nor worsens algorithmic disparities in a systematic way, and that fairness auditing and, where necessary, mitigation remain necessary regardless of the model class.

The CMES condensed these competing profiles into a single score, but the resulting differences were modest (mean CMES between 0.855 and 0.881) and were partly compensatory, as the weakness of the foundation models in robustness was offset by their strengths in calibration and predictive performance. The statistical analysis supports a moderately strong conclusion: the omnibus test was significant (Table 6), and TabFM, TabICL, and TabDPT were each significantly better than Logistic Regression, with TabDPT also significantly better than XGBoost (Table 8). However, the foundation models were statistically indistinguishable from each other, and TabPFN could not be separated from either the best foundation models or the baselines (Figure 7). The ranking within the top group should therefore not be over-interpreted, particularly because the best model changed from one dataset to another (Figure 1 (C) and Figure 7 (C)).

### 3.9 Limitations and Future Work

This study has several limitations. First, majority-class downsampling was applied to datasets exceeding 700 observations to establish a constrained small-data benchmarking setting. However, this altered the natural class distributions and disease prevalence, potentially affecting predictive values, calibration, fairness, and model behavior under real-world conditions. Both the balanced datasets used in this study and their original imbalanced versions are available in our GitHub repository. Future research should extend this evaluation to the full datasets with their natural class distributions to assess model performance under realistic prevalence and varying degrees of class imbalance.

Second, the use of ten public datasets and internal five-fold cross-validation may limit the generalizability of our findings to other populations and clinical settings. External, temporal, and prospective validation using larger and more diverse clinical cohorts is therefore warranted to establish the reliability of these models beyond the evaluated datasets.

Third, fairness evaluation was restricted to binary sex-based comparisons, while robustness assessment relied on Gaussian feature perturbations. These analyses do not capture broader demographic disparities, intersectional inequities, or other clinically relevant sources of data variability. Future studies should incorporate intersectional fairness analyses and more realistic perturbation and distribution-shift scenarios, alongside investigations of potential fairness and robustness mitigation strategies.

Finally, the CMES depends on the selected metrics and their weighting, potentially obscuring dimension-specific trade-offs. Future work should examine alternative weighting schemes and incorporate clinical utility and decision-analytic outcomes to determine whether benchmark performance translates into meaningful clinical benefit.

## 4 Conclusion

This study presents a multidimensional benchmark of four tabular foundation AI models, TabFM, TabPFN, TabDPT, and TabICL, against Logistic Regression and XGBoost across ten WHO-aligned healthcare classification datasets. By evaluating predictive discrimination, sex-based fairness, probabilistic calibration, and robustness under controlled Gaussian feature perturbations within a standardized cross-validation framework, this study provides a comparative assessment of the trustworthiness-related properties of contemporary tabular models in healthcare. Overall, the foundation models demonstrated favorable predictive and probabilistic performance relative to the conventional baselines. TabFM achieved the highest mean CMES, followed closely by TabDPT, TabICL, and TabPFN. It also attained the highest mean AUROC and sensitivity, together with the lowest ECE, NLL, and Brier score, suggesting comparatively strong discrimination and probability reliability across the evaluated datasets. Nevertheless, the statistically significant omnibus comparison did not translate into significant pairwise differences among the four foundation models after multiple-comparison correction, indicating that their overall performance differences were relatively small and dataset-dependent. In conclusion, TFMs offer promising predictive and calibration advantages for healthcare classification under constrained data conditions, but these advantages are not universal across trustworthiness dimensions. Their responsible use requires context-specific evaluation, transparent reporting, and validation beyond benchmark performance. The proposed framework provides a reproducible foundation for such evaluations and supports more evidence-informed selection of tabular models for future digital health research.

## Data Availability

All data and code underlying the findings of this study are publicly available without restriction in the study’s GitHub repository (https://github.com/e1research/tfm-healthcare). The repository further provides source links for all ten original publicly available clinical datasets, the harmonized, standardized, and downsampled benchmark datasets evaluated in this study, and additional supplementary dataset variants, including versions preserving the original sample sizes and natural class prevalences.

